# Personalized prediction of UTI risk and antibiotic susceptibility based on gut *E. coli*

**DOI:** 10.1101/2024.04.05.24305377

**Authors:** Veronika Tchesnokova, Debarati Choudhury, Lydia Larson, Irina Basova, Yulia Sledneva, Teresa Cristina Bonilla, Thalia Solyanik, Jennifer Heng, Isaac Pasumansky, Victoria Bowers, Sophia Pham, Julia Seregin, Gregory S. Davis, Lawrence T. Madziwa, Erika Holden, Sara Y. Tartof, James D. Ralston, Evgeni V. Sokurenko

## Abstract

Community-acquired UTI is the most common bacterial infection. We assessed whether gut *Escherichia coli* profiles are associated with UTI risk and pathogen antibiotic susceptibility in women aged ≥50, antibiotics-free for ≥1 year. In an 18-month prospective observational study, 1,804 participants provided a baseline fecal sample screened for *E. coli* abundance, resistance to trimethoprim/sulfamethoxazole, ciprofloxacin, and third-generation cephalosporins, and association with pandemic multidrug-resistant (MDR) clonal groups ST131-*H30* and ST1193. *E. coli* was present in 90.8% of samples; 37.5% showed resistance to one or more antibiotics. MDR strains were highly abundant in the gut, with ST131-*H30* carriage increasing and ST1193 declining with age. During the 18-months follow-up, UTIs occurred in 10.9% of *E. coli* carriers versus 3.3% of non-carriers (P=.003), with the highest risk of 37.8% observed among women ≥70 yo colonized with ST131- *H30*. Increased fecal abundance of strains resistant to the antibiotics strongly associated with the UTI risk. The original fecal *E. coli* resistance profiles were concordant with UTI antibiotic susceptibility with 95–100% accuracy in 95% of patients.

## Introduction

The personalized or precision medicine approach aims to prospectively define disease risks, expected outcomes, and optimal treatments based on an individual’s unique biomarkers, usually determined genetically but also reflecting behavioral factors^1^. The indigenous microbiota of the gut, vagina, and other body compartments is considered an integral part of the human organism, influencing many aspects of health^2,3^. At the same time, colonizing microbiota serves as a major reservoir of opportunistic pathogens, including strains resistant to antibiotics^4–6^. However, it remains unclear to what extent identifying specific colonizers species, phylogenetic lineages and the resistance profiles can predict future infection probability and optimal antibiotic treatment – a critically important question in the era of rising antimicrobial resistance, particularly in individuals at risk for severe outcomes.

Gut colonizing *Escherichia coli* as well as other enterobacteria, are the source of strains causing community-acquired urinary tract infections (UTIs) – the most common bacterial infection treated in primary care worldwide^7–10^. Postmenopausal women, typically aged 50 and older, are at especially high risk of severe and drug-resistant UTIs^11,12^. Acute uncomplicated cystitis is the most frequent presentation, imposing a substantial economic burden, especially when initial antibiotic treatment fails^10,13^. Complicated UTI, recurrent cystitis and severe UTI forms (like pyelonephritis and urosepsis) – further increase costs and morbidity, particularly in the elderly^10,13,14^.

UTIs represent a leading cause of antibiotic use in humans^15^. Because culture-based susceptibility testing takes 2-4 days, clinicians typically rely on empiric (pre-culture) therapy. This approach is inherently imprecise, as it is based on community-level resistance trends rather than the resistance profile of the infecting strain^16^. For empiric treatment of uncomplicated UTI, the Infectious Diseases Society of America (IDSA) recommends antibiotics only if the local *E. coli* resistance prevalence is ≤20%, while for severe infections the threshold is <10%^16–18^.

While *E. coli* strains are generally susceptible to nitrofurantoin and fosfomycin, these agents are not appropriate for individuals with reduced renal function or at risk for severe UTIs – conditions that are common in older adults^19,20^. Resistance among *E. coli* to other commonly used UTI antibiotics – such as trimethoprim/sulfamethoxazole (TMP/SMX) or ciprofloxacin (CIP) – continues to rise fast, commonly exceeding recommended thresholds and driving overuse of broader-spectrum agents, like third-generation cephalosporins (3GCs). This trend contributes to increased mortality, healthcare expenditures and emergence of resistance to last-line antibiotics. Each ineffective antibiotic prescription (“drug-bug mismatch”) is on average associated with approximately $2,000 in additional cost^21–24^. Particularly concerning is the global emergence of multidrug-resistant (MDR) *E. coli* strains, defined as resistant to three or more antimicrobial classes. Two MDR clonal groups dominate worldwide: ST131- *H30* (aka clade C1/C2) and ST1193^25–32^. Both lineages are distinguished by hallmark fluoroquinolone resistance and rose to pandemic prominence in the early 2000s and 2010s, respectively.

It was shown that gut carriage of MDR *E. coli* increases the risk of extraintestinal infections, including UTIs, in hospitalized patients, transplant recipients, and preterm infants^33–38^. However, existing studies are limited, short-term (0.5–3 months), and focused on highly selected populations. To our knowledge, no study has prospectively assessed whether gut-colonizing bacterial profiles can predict UTI risk or uropathogen antibiotic susceptibility in the general community over relatively long time periods.

Here, we conducted a prospective observational study of non-hospitalized women aged >50 years, with no antibiotic use or UTI in the prior year. We evaluated whether profiling gut-colonizing *E. coli* strains could support a personalized-medicine approach for managing community-acquired UTIs, particularly in at-risk patients, enhance antimicrobial stewardship efforts, and strengthen the rationale for developing decolonization or other strategies to prevent severe infections^33,39–42^.

## RESULTS

### Resistant *E. coli* are carried by one-third of women with contrasting age-prevalence of CIP-R strains

We collected fecal samples from 1,804 study enrollees – 437 women (24%) aged 50-59, 632 (35%) aged 60-69, 532 (30%) aged 70-79, and 203 (11%) aged ≥80 (**Supplemental Figure S1A**).

Fourteen samples (0.8%) failed to yield any visible bacterial growth on the chromogenic UTI agar plates, while *E. coli* was detected in 1,638 (90.8%) (**Figure 1A**), often in mixture with other species (see below). In 8.5% of samples only non-*E. coli* bacteria grew, primarily other Gram-negative species (e.g., *Klebsiella, Enterobacter, Citrobacter*) or Gram-positive species (mainly *Enterococcus*). The overall prevalence of *E. coli*-positive samples was slightly higher in women aged ≥70 (OR=1.75, 95% CI: 1.22-2.62, P=.0017) (**Figure 1B)**.

**Figure 1.**
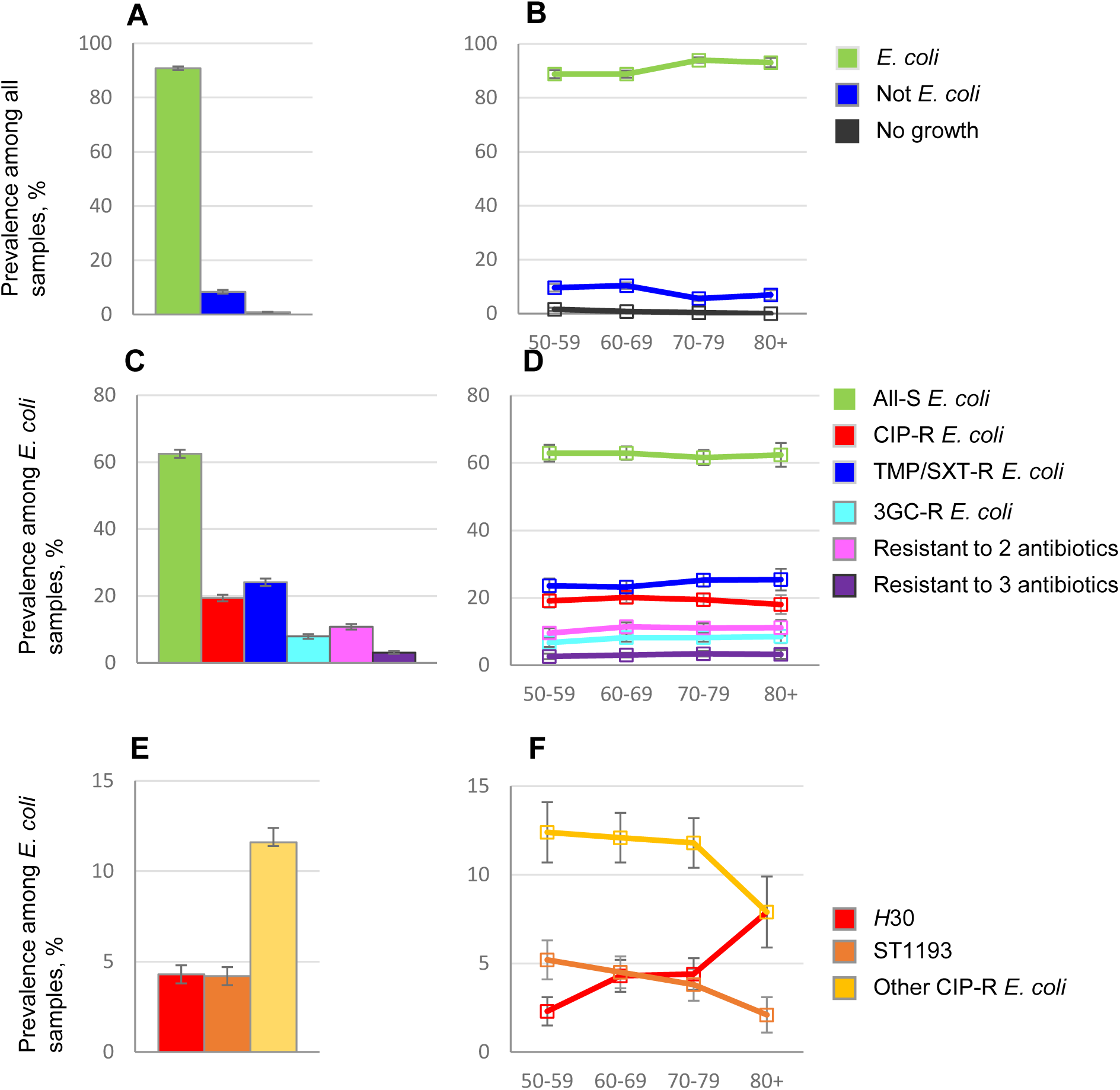
Total prevalence of fecal samples carrying different categories of *E. coli* strains. Prevalence of samples with and without any *E. coli* growth across all ages (A) and in different age groups (B); Prevalence of samples yielding the growth of *E. coli* on different antibiotics across all ages (C) and in different age groups (D); Prevalence of samples containing *E. coli* from ST131-H30, ST1193 and combined minor clonal groups across all ages (E) and in different age groups (F). Error bars represent SEM.

In 62.5% of the *E. coli*-positive samples, no *E. coli* resistant to either TMP/SXT, CIP, or 3GC were recovered (‘all-sensitive’ samples), while the rest contained *E. coli*resistant to at least one of the antibiotics: 24.1% to TMP/SXT, 19.4% to CIP, and 7.9% to 3GC (**Figure 1C**). In 10.8% and 3.1% of samples, *E. coli* were resistant to two or all three antibiotics, respectively (**Figures 1C-D and Supplemental Figure S2**). There were no major age differences in the carriage of *E. coli* with different resistance profiles (**Figure 1D**) (**Supplemental Figure S1B-E**). Among the 177 samples with growth on multiple antibiotics, in 122 (68.9%) samples multiple resistance was attributable to a single strain, based on a high-resolution *fimC/fimH* clonal profiling (*clonotyping* - see Methods), with 50 strains resistant to all three antibiotics (i.e., MDR strains). The remaining 55 samples yielded a mixture of multiple resistant strains.

The CIP-resistant (CIP-R) strains dominated among isolates with multiple resistance (92.5% of 157 strains, P<.001; **Supplemental Figure S2**). Clonotyping revealed that nearly half (43.4%) of CIP-R *E. coli* belonged to two globally disseminated clonal groups: ST131-*H30* (22.0%) and ST1193 (21.4%) (**Figure 1E**), which also dominated among MDR strains (**Supplemental Table S1**). The remaining CIP-R isolates were distributed among 113 minor clonotypes, each far less common than the pandemic lineages (**Supplemental Table S1**). ST131-*H30* was more common among women aged ≥80 than 50-59 (7.9% vs. 2.3%, respectively; OR=5.70, 95% CI: 1.95-17.0, P=.0002), while both ST1193 and minor clonal groups combined were more common in the youngest than oldest group (16.8% vs. 10.1%, respectively) (**Figure 1F**). The opposite age trends between ST131-*H30* and other CIP-R *E. coli* were further evident in the age-stratified analyses **(Suppl. Figure S1F-G).**

### *E. coli* is the predominant uropathogen in the gut

The chromogenic UTI agar is designed to differentiate *E. coli* from other Gram-negative and Gram-positive uropathogens. Fecal samples were plated using the four-quadrant streak method (examples in **Supplemental Figure S3A**) – a standard approach for (semi)quantitative assessment of the bacterial abundance. By using 16S metagenomics, we validated that four-quadrant plating on UTI agar provides an effective estimate of the relative abundance of *E. coli* vs. other bacterial species (see Materials and Methods and **Supplemental Figure S3C**).

Among the *E. coli*-positive samples, *E. coli* completely or greatly dominated over other Gram-negative bacteria (**Figure 2**). Nearly 70% of samples contained *E. coli* at ≥90% abundance relative to all Gram-negative bacteria. Other commonly identified Gram-negative bacteria included various *Klebsiella* or *Enterobacter* species. Relative to total bacterial growth on UTI agar, *E. coli* abundance was also high but lower than against Gram-negative bacteria alone. Here, the distribution was distinctly bimodal (**Figure 2**) and included a cluster of samples where *E. coli* accounted for roughly half of colonies, with the rest being primarily enterococci species. *E. coli* abundance patterns were consistent across age groups (**Supplemental Figure S4A-B**).

**Figure 2.**
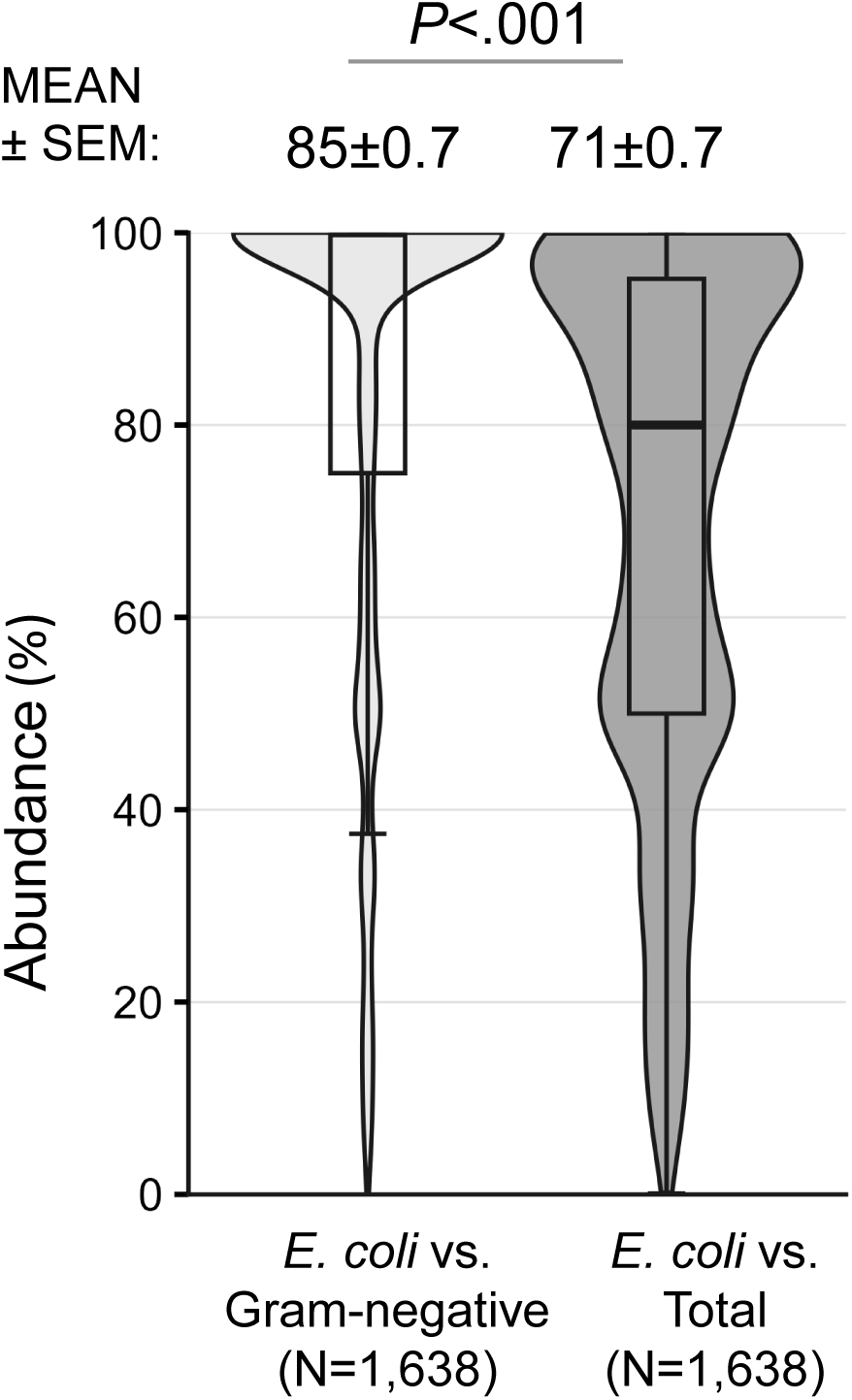
Combined violin/box plot distribution of *E. coli* abundances in fecal samples compared with Gram-negative bacteria and total bacteria grown on UTI agar. Sample sizes for each group are shown in parentheses. Average abundances (Mean ± SEM %) are displayed above the plots. Above the solid line, *P* values between the average abundances are displaced for only statistically significant differences.

### CIP-R *E. coli* strains from pandemic clonal groups are highly prolific gut colonizers

We assessed the relative abundance of CIP-R *E. coli* versus other *E. coli* in the gut by comparing growth on UTI agar plates with and without CIP. All CIP-R *E. coli* strains combined showed a bimodal distribution (**Figure 3A**). Most resistant *E. coli* strains were either dominant (≥90%) or rare (≤10%) within the gut, with the latter pattern being more common overall (28% vs. 38%, P=.011). However, strains from ST131-*H30* and ST1193 were twice as likely to occur as dominant rather than rare sub-populations: 27% vs. 14% (P=.061) and 35% vs. 17% (P=.019), respectively (**Figure 3A**). In contrast, the combined minor CIP-R clonotypes were twice *less* likely to occur as the dominant rather than the rare sub-population (25% vs. 54%, P<.001). The relative prevalence of CIP-R strains did not correlate with *E. coli* abundance relative to Gram-negative or total bacteria grown on UTI agar (**Supplemental Figure S4C-D**).

**Figure 3.**
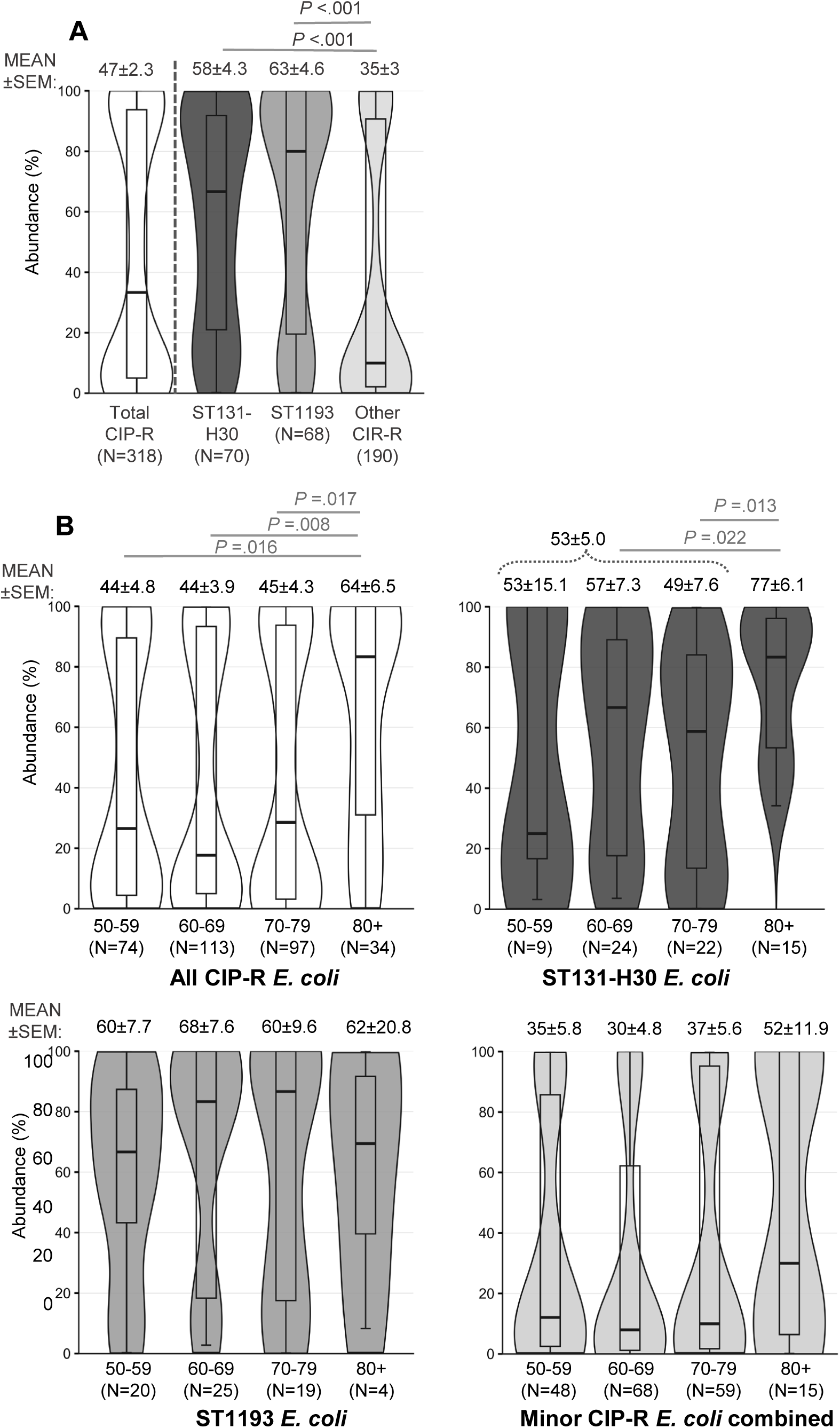
Combined violin/box plot of abundance of CIP-R *E. coli* relative to total *E. coli* in fecal samples across all ages combined (A) and within each age group (B). Sample sizes for each group are shown in parentheses. Average abundances (Mean ± SEM %) are displayed above the plots (For ST131-H30, also displaced, are the average values for ages <80 combined). Above the solid line, *P* values between the average abundances are displaced for only statistically significant differences.

Age-wise, CIP-R *E. coli* were significantly more abundant in women aged ≥80 compared with younger women (medians 83.3% vs. 25%, respectively, P=.011) (**Figure 3B**), primarily due to the higher abundance among the elderly of ST131-*H30*. Both ST131-*H30* and ST1193 were generally more abundant than other clonotypes across all age groups, though differences diminished in women aged ≥80.

### UTI risk is associated with *E. coli* gut carriage, multidrug resistance, ST131-*H30*, and older age

Based on the combinatorial criteria for the clinical UTI^7^, 184 women (10.3%) experienced at least one UTI episode during the 18 months of follow-up observation (**Supplemental Figure S5, Table S5**), all in the outpatient setting (hereafter, the diagnosed UTI cohort). UTIs occurred about three times more frequently in women with detectable gut *E. coli* compared to those without - 10.9% vs. 3.3%, respectively (OR=8.8, 95% CI: 1.5-11.4, P=.003) (**Figure 4A**). UTI risk did not differ significantly between those colonized with susceptible strains or resistant strains (OR=1.12, 95% CI 0.80-1.58, P=.480), though women colonized with MDR *E. coli* were more likely to present with UTI than the rest of women - 18.0% (9/50) vs 9.5% (54/566), respectively (OR=2.08, 95% CI: 0.84-4.75, P=.059) (**Figure 4C**).

**Figure 4.**
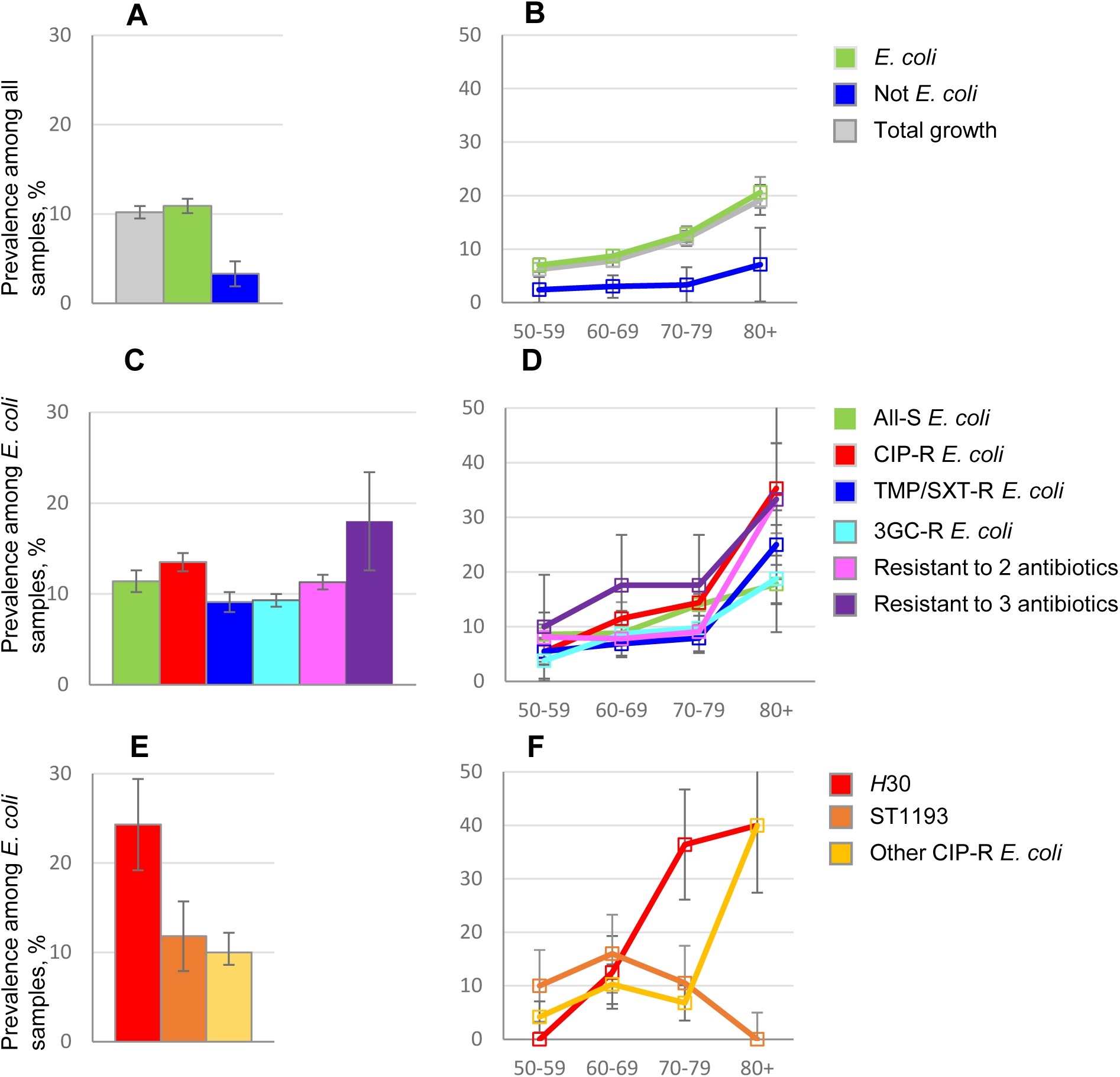
Dependence of UTI frequencies on the characteristics of colonizing *E. coli*. UTI frequencies of *E. coli* carriers and non-carriers across all ages (A) and in different age groups (B); UTI frequencies in carriers of *E. coli* with different resistance profiles across all ages (C) and in different age groups (D); UTI frequencies among carriers of different clonal groups associated with CIP-R *E. coli* across all ages (E) and in different age groups (F). Error bars represent the standard error. UTI frequencies are based on the diagnosed UTI cohort (n=184)

While carriers of ST1193 or minor CIP-R clonotypes experienced similar numbers of infections, those colonized with ST131-*H30* experienced significantly more infections - 24.3% (17/70, OR=3.01, 95% CI: 1.59-5.42, P=.0001) (**Figure 4E**). Notably, 5 of the 9 MDR-associated UTIs occurred in women colonized with ST131-*H30*. When stratified by age, UTI risk increased with advancing age, reaching 19.2% among women aged ≥80 years (50/189, OR=2.46, 95% CI: 1.63-3.66, P<.0001) (**Figure 4B**). This age dependence was observed in both *E. coli* carriers and non-carriers (**Figure 4B**), and across different resistance profiles (**Figure 4D** and **Supplemental Figure S1I-M**). The most pronounced increase, however, was in women ≥80 years colonized with CIP-R or MDR *E. coli*, where rates exceeded 30%. The strongest age-associated risk was among carriers of ST131-*H30*, with a four-fold increase in women ≥70 years relative to the younger study enrollees – 37.8% (14/37) vs 9.1% (3/33), respectively (OR=6.1, 95% CI 1.4-36.0, P = .0051) (**Figure 4F**).

### Vast majority of antibiotic-resistant UTIs originates from the baseline fecal *E. coli*

According to the EHR, among urine samples submitted for analysis to the clinical microbiology laboratory, 134 had uropathogens with defined antimicrobial susceptibility profiles (**Supplemental Figure S4C-D**) (hereafter, the pathogen-defined UTI subgroup). Of the 134 pathogen-defined UTI cases, 118 (88.1%) were caused by *E. coli* and 16 by non-*E. coli*pathogens (see below). Among uropathogenic *E. coli*, 82 (69.5%) were susceptible to all three tested antibiotics, 19 (16.1%) resistant to TMP/SXT, 19 (16.1%) to CIP, and 5 (4.2%) to 3GC (**Supplemental Table S2**). Among the 16 non-*E. coli* UTI cases, uropathogens included *Klebsiella species* (9), *Proteus mirabilis* (3), *Citrobacter species* (2), *Salmonella enterica* (1), and *Enterococcus faecalis* (1). In contrast to *E. coli*, all non-*E. coli* uropathogens were fully susceptible to the three antibiotics tested (**Supplemental Table S3**).

All participants with *E. coli*-positive UTIs had *E. coli* in their baseline fecal samples. When the urinary isolate was fully susceptible, 84.1% (69 of 82) of the corresponding fecal samples yielded only all-susceptible *E. coli.* Of the 36 urinary isolates resistant to one or multiple antibiotics, 31 (86.1%) had a matching resistance profile in the baseline feces, with concordance rates of 89.5% for resistance to either TMP/SXT or CIP and 80% for resistance to 3GC. Five urine samples contained resistant pathogens that did not match the fecal *E. coli* profiles, with those UTIs occurring on months 4, 6, 13, 16, and 17 of the observation (**Supplemental Figure S5; Supplemental Table S2**).

A total of 76 *E. coli*-positive urine samples were obtained from the clinical microbiology laboratory and analyzed in detail. Each sample was plated, and five colonies were randomly selected for clonotyping. In 75 (98.7%) samples, *E. coli* colonies were clonally homogeneous, suggesting a single dominant strain. Shotgun metagenomic sequencing from 35 randomly selected urine samples confirmed 100% abundance of a single strain, with a detection limit of ∼1%. In the single clonally heterogeneous urine sample, metagenomic analysis identified two distinct *E. coli* strains in similar proportions belonging to the clonal groups ST200 and ST141.

Of the 76 urinary *E. coli* isolates analyzed, 25 were resistant to at least one antibiotic. Among these, 22 (88.0%) had matching both resistance and clonal profiles with the corresponding same-person fecal isolates. In the three cases with non-matching resistance profiles, urine and fecal isolates belonged to different clonal identities (**Supplemental Data**). Among the remaining 51 all-susceptible urinary isolates, a clonally identical strain was found by culture-based methods in 23 (40.1%) of the corresponding baseline fecal samples, including the single clonally heterogeneous case. Metagenomic sequencing did not identify any additional fecal samples with all-susceptible strains clonally matching the same-person urinary isolates.

Whole-genome sequencing was successfully performed on 42 of the 45 clonally matched pairs. Across 2,283 core genes, urinary and fecal strains from the same individual differed by only 3.2 ± 0.6 SNPs on average (**Supplemental Figure S6**). Irrespective of clonotype, same-person fecal- urinary isolate pairs consistently clustered together on the phylogenetic tree, distinct from the closest publicly available genomes of the same ST **(Supplemental Figure S6)**, strongly suggesting that the paired isolates originated from the same strain. All 16 participants with non-*E. coli*-positive UTIs also had *E. coli* in their baseline fecal samples, with *E. coli* resistant to TMP/SXT present in 2 samples and to CIP in 1 sample (**Supplemental Table S3**). No in-depth search for these uropathogens was performed in the corresponding fecal samples.

### UTI-causing *E. coli* strains are found in feces at higher abundance

The abundance of total gut *E. coli* relative to Gram-negative bacteria or UTI agar-grown bacteria did not differ between women who did or did not develop UTI (**Figure 5A-B**). However, in women who experienced a UTI caused by a resistant *E. coli*, the relative gut abundance of the pathogen was higher than in UTI-free carriers of resistant *E. coli* (**Figure 5C**). A similar pattern was also observed when analyzing CIP-R, TMP/SXT-R, and 3GC-R *E. coli* separately, as well as among different CIP-R clonal groups (**Supplemental Figure S7**).

**Figure 5.**
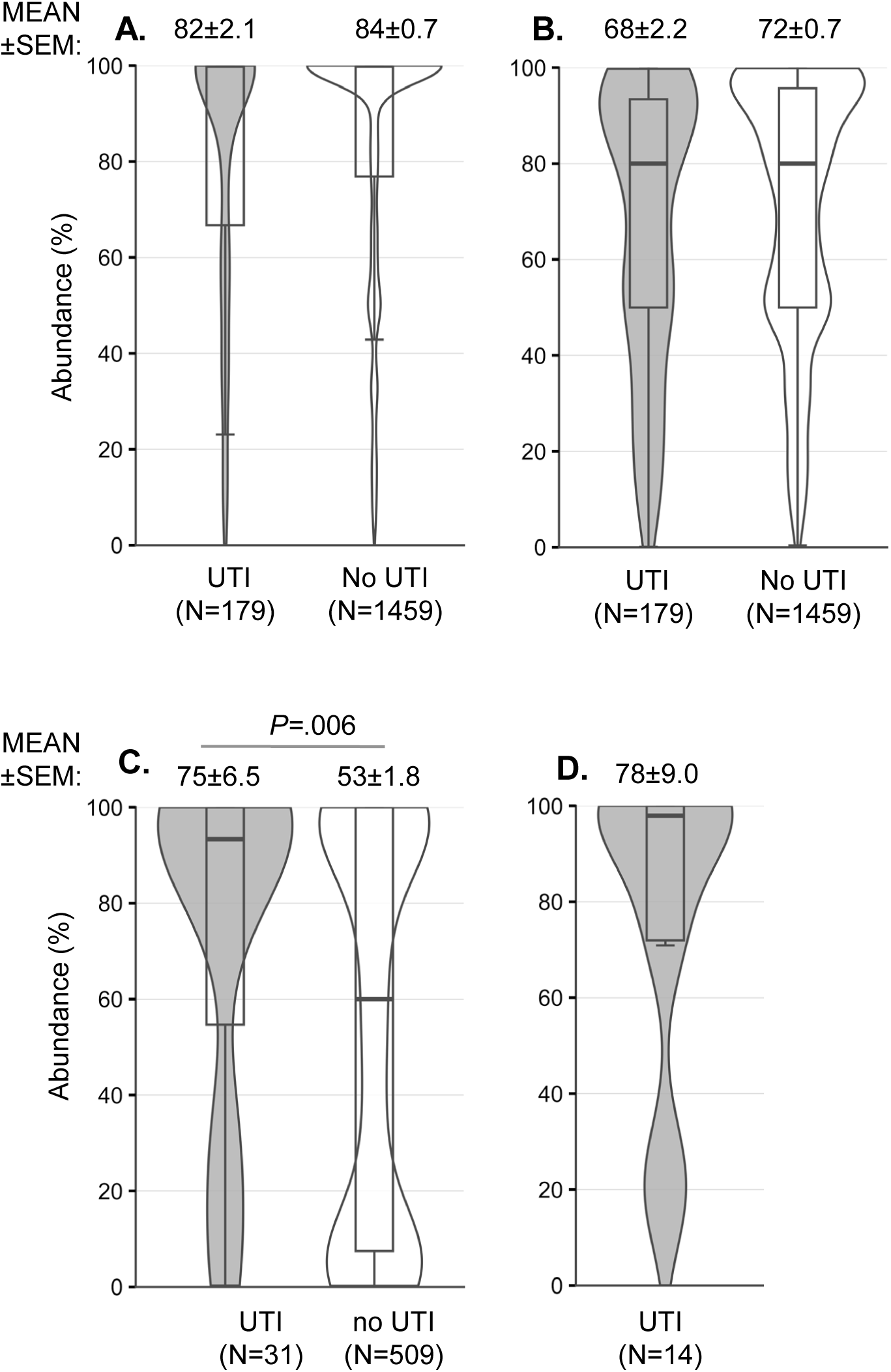
**Violin/box plot of *E. coli* abundances in fecal samples of women with and without UTI. A-B**. For all *E. coli*-positive samples with determined abundance of *E. coli* vs. Gram-negative bacteria (**A**) or vs total bacteria grown on the UTI agar (**B**). (**C**) For any resistant *E. coli*-positive samples the abundance of uropathogenic resistant *E. coli* vs. total *E. coli* (UTI cohort includes only UTI caused by matching resistant *E. coli*). (**D**) For cohort of UTI caused by all-sensitive *E. coli*, abundance of uropathogen vs. total *E. coli*. Sample sizes for each group are shown in parentheses. Average abundances (Mean ± SEM %) are displayed above the plots. Violins are scaled by area. Above the solid line, *P* values between the average abundances are displaced with only statistically significant differences.

Women carrying CIP-R gut *E. coli* as a subpopulation at >50% abundance had a threefold higher probability of developing a CIP-R UTI compared with those carrying CIP-R strains at ≤50% abundance (OR=3.23, 95% CI: 1.03–12.50, P = .0229). The probability increased further among women carrying CIP-R strains at ≥90% abundance compared with those at ≤10% (OR=7.69, 95% CI: 0.70–333.34, P = .0471).

Metagenomic analysis of 36 fecal samples revealed an average of 1.77±0.12 (range 1-4) *E. coli* CH clonotypes per sample (**Supplemental Table S5, Supplemental Methods**). In the subset of 14 samples from women whose UTIs were caused by all-sensitive *E. coli* that clonally matched the same-person fecal isolates, based on both metagenomic and single-colony analyses, which showed high correlation (**Supplemental Figure S8**), the all-sensitive UTI strains also tended to represent dominant fecal subpopulations (**Figure 5D**). To directly assess whether clonal diversity influenced UTI risk, we compared *E. coli* CH clonotype richness in ST131-*H*30-carrying fecal samples between women who did and did not develop a UTI during follow-up using amplicon sequencing (**Supplemental Table S5)**. The number of clonotypes per sample did not differ significantly between the two groups (1.93 ± 0.18 vs. 1.64 ± 0.17, mean ± SEM; P=.254), suggesting that it is the presence of specific high-risk clones rather than overall clonal diversity that drives UTI susceptibility. Additionally, we estimated the number of distinct *E. coli* CH clonotypes within fecal samples for a subset of 20 cases in which a UTI episode occurred and the fecal sample carried resistant *E. coli*. There was a non-significant trend towards a lower clonotype count in samples where the resistant clone went on to cause a UTI compared to those where it did not (N=10 each group, 1.9 ± 0.4 vs. 2.4 ± 0.3, mean ± SEM; P=.295). No analysis was performed for women without UTI who carried all-sensitive strains of specific clonotypes.

### Antibiotic susceptibility of UTI pathogens can be predicted by testing the fecal *E. coli*

We evaluated how accurately the susceptibility of clinical urine isolates could be predicted, i.e., the antibiotic choice is advisable, by testing the susceptibility of *E. coli* in prospectively collected fecal samples. This analysis was restricted to the *pathogen-defined* UTI subgroup (n=134), which included both *E. coli*-caused UTIs (n=118) and non-*E. coli* UTIs (n=16), all of the latter being fully susceptible to the three antibiotics tested (see above). Overall, in our study, 85.1% of uropathogens were susceptible to TMP/SXT, 85.8% to CIP, and 95.5% to 3GC; 95.5% were susceptible to either TMP/SXT or CIP, and 98.5% to at least one antibiotic (Table 1).

**Table 1.**
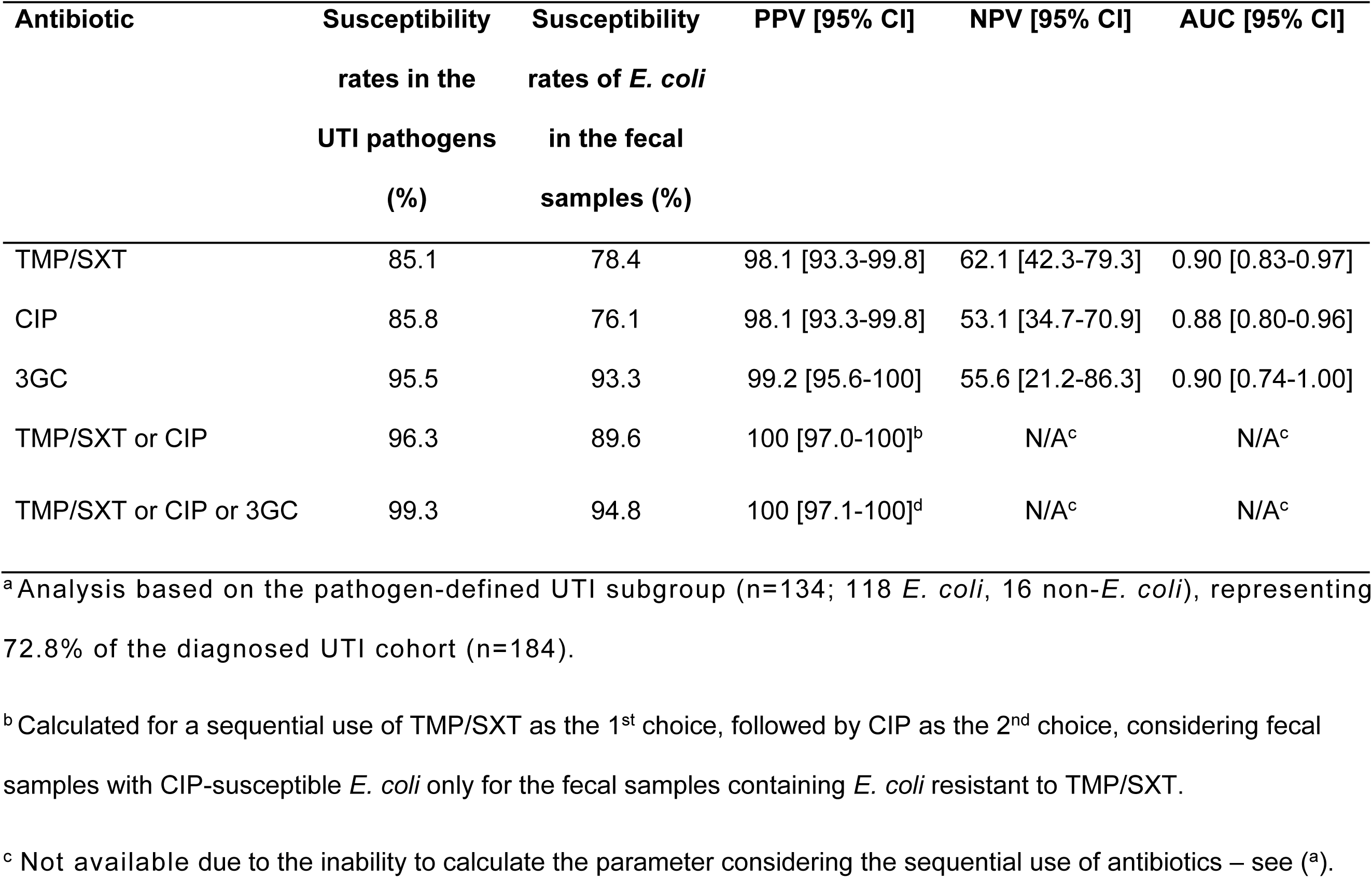
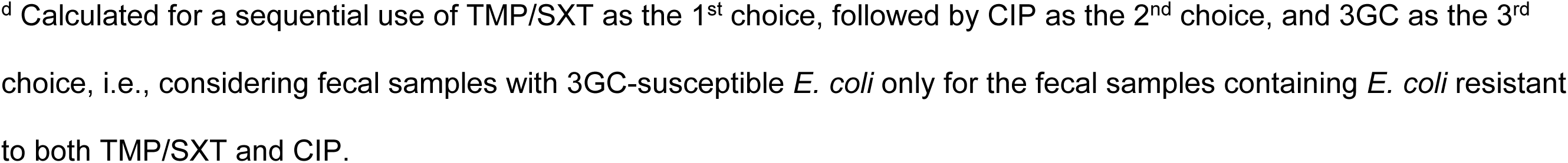
Susceptibility prediction of the UTI pathogens based on fecal *E*.

| Antibiotic | Susceptibility<br>rates in the<br>UTI pathogens<br>(%) | Susceptibility<br>rates of *E. coli*<br>in the fecal<br>samples (%) | PPV [95% CI] | NPV [95% CI] | AUC [95% CI] |
| --- | --- | --- | --- | --- | --- |
| TMP/SXT | 85.1 | 78.4 | 98.1 [93.3-99.8] | 62.1 [42.3-79.3] | 0.90 [0.83-0.97] |
| CIP | 85.8 | 76.1 | 98.1 [93.3-99.8] | 53.1 [34.7-70.9] | 0.88 [0.80-0.96] |
| 3GC | 95.5 | 93.3 | 99.2 [95.6-100] | 55.6 [21.2-86.3] | 0.90 [0.74-1.00] |
| TMP/SXT or CIP | 96.3 | 89.6 | 100 [97.0-100]<sup>b</sup> | N/A<sup>c</sup> | N/A<sup>c</sup> |
| TMP/SXT or CIP or 3GC | 99.3 | 94.8 | 100 [97.1-100]<sup>d</sup> | N/A<sup>c</sup> | N/A<sup>c</sup> |

<sup>a</sup> Analysis based on the pathogen-defined UTI subgroup (n=134; 118 *E. coli*, 16 non-*E. coli*), representing 72.8% of the diagnosed UTI cohort (n=184).
<sup>b</sup> Calculated for a sequential use of TMP/SXT as the 1<sup>st</sup> choice, followed by CIP as the 2<sup>nd</sup> choice, considering fecal samples with CIP-susceptible *E. coli* only for the fecal samples containing *E. coli* resistant to TMP/SXT.
<sup>c</sup> Not available due to the inability to calculate the parameter considering the sequential use of antibiotics – see (<sup>a</sup>).
<sup>d</sup> Calculated for a sequential use of TMP/SXT as the 1<sup>st</sup> choice, followed by CIP as the 2<sup>nd</sup> choice, and 3GC as the 3<sup>rd</sup> choice, i.e., considering fecal samples with 3GC-susceptible *E. coli* only for the fecal samples containing *E. coli* resistant to both TMP/SXT and CIP.

As mentioned above, *E. coli* was present in all corresponding baseline fecal samples. In 78.4% of fecal samples, *E. coli* was susceptible to TMP/SXT, in 76.1% to CIP, and in 93.3% to 3GC; in 89.6% of samples, *E. coli* was susceptible to either TMP/SXT or CIP, and in 94.8% to at least one antibiotic (Table 1). The performance of the fecal testing was first evaluated without considering the relative abundance of resistant *E. coli* in the baseline samples.

We estimated the probability that the urinary pathogen would be susceptible to a specific antibiotic based on the absence of resistant *E. coli* in the fecal sample – i.e., the test’s positive predictive value (PPV), calculated as the ratio of correct susceptibility predictions (true positives) to the sum of true and (incorrectly-predicted) false positives. The PPV values for susceptibility to individual antibiotics were higher than 98% (Table 1). Thus, in only a very small number of the antibiotic-advisable portion of UTI cases (from 76.1% for CIP to 93.3% for 3GC) would fecal *E. coli* testing incorrectly predict that the uropathogen is susceptible to a given antibiotic. Consequently, a similar level of accuracy was maintained when considering susceptibility to a sequential use of TMP/SXT and then, CIP, which together would cover nearly 90% of UTI cases, while susceptibility to 3GC could be accurately predicted in approximately half of the remaining 10% cases.

We also assessed the probability that the urinary pathogen would not be susceptible to a specific antibiotic based on the presence of resistant (non-susceptible) *E. coli* in the fecal sample – i.e., the test’s negative predictive value (NPV), calculated as the ratio of correct predictions of non-susceptibility (true negatives) to the sum of true and false negatives. The NPV values ranged from 53.1% for CIP to 62.1% for TMP/SXT (Table 1). Although in a substantial number of cases the pathogen would be mistakenly predicted being resistant, using fecal testing would still prevent a very high rate of drug–bug mismatch in the antibiotic-inadvisable portion of UTI cases (ranging from 23.9% for CIP to 6.7% for 3GC).

To investigate the basis of discordant resistance predictions, we performed clonal and plasmid analysis on 20 discordant fecal-urinary *E. coli* pairs available for typing. Most discordant cases (16/20) were explained by either acquisition of a new uropathogenic clone not detected in the baseline fecal sample, or by the presence of multiple co-colonizing *E. coli* strains with different resistance profiles in the fecal sample, only one of which caused the UTI. In the remaining 4 cases, the UTI was caused by a strain clonally matched to the fecal isolate that had lost some of its resistance. Plasmid analysis identified the mechanism in one of these cases (ID10226), in which the urinary ST1193 isolate had lost an IncI1-I(Alpha) plasmid carrying *bla*CTX-M-55 relative to its fecal counterpart. In the remaining three cases (ID16572, ID15727, and ID16660) – involving CIP-R uropathogens ST69, ST450, and ST131-*H*30, respectively – the exact mechanism underlying resistance loss could not be determined from short-read whole-genome sequencing data.

Based on the obtained NPV and PPV values, we then calculated the area under the ROC curve (AUC) that could serve as a summary metric for the potential diagnostic performance of the fecal test in guiding empiric (pre-culture) antibiotic selection. For the three antibiotics, AUC values ranged from 0.88 to 0.90 (Table 1), indicating excellent predictive performance43.

Finally, we determined that, by applying different cutoffs for the fecal abundance of resistant *E. coli*, the prediction of uropathogen non-susceptibility to CIP and 3GC (NPV values) could be notably improved (Supplemental Figure S9). Although this was accompanied by a reduction in susceptibility prediction accuracy (PPV values), the overall diagnostic performance of the fecal test, as indicated by the corresponding AUC values, remained high.

## Discussion

Our study demonstrates that both the presence and clonal identity of gut *E. coli* influence the risk of community-acquired UTI within at least 18 months, and that defining the resistance profiles of gut *E. coli* shows potential as a pre-culture predictive tool for uropathogen susceptibility. While our findings are proof-of-concept and primarily aimed at establishing the feasibility of fecal testing for personalized UTI risk stratification and treatment guidance, such testing could support a more individualized approach to UTI management by improving infection risk prediction and guiding empiric antibiotic selection, which may be particularly valuable in at-risk individuals. This conclusion was reached based on the study of women aged 50 and older, who are typically considered pre- to postmenopausal and prone to UTIs due to estrogen decline, higher vaginal pH, increased bladder urine retention, and other factors^43,44^. Because our study enrollees supposedly had neither antibiotic exposure nor UTI in the year before enrollment and, thus, may have produced a lower-risk cohort, the potential impact of fecal *E. coli* testing may prove even greater in populations susceptible to more frequent and/or severe infections and higher risk of acquiring MDR pathogens.

As expected, *E. coli* was isolated from fecal samples in most women. Interestingly, women with no fecal *E. coli* recovered were a minority for yet unexplained reasons, *E. coli* non-carriers had approximately one-third the risk of developing UTIs compared to carriers, indicating a strong association between *E. coli* presence and UTI incidence. Due to the fecal sample collection setup, we could not measure the *E. coli*abundance in the entire fecal microbiome as done previously^35^. We show, however, that *E. coli* is more abundant in the gut relative to other known uropathogens. Moreover, we show that strains causing UTI tend to represent the most abundant *E. coli* sub-population in the gut, supporting the hypothesis that *E. coli* fitness as a gut colonizer is closely linked to its success as a urinary tract pathogen^8,45–48^.

In our study, we focused on three antibiotics commonly used for community-onset UTI. TMP/SXT is a recommended first-line treatment for uncomplicated UTI, along with nitrofurantoin or fosfomycin^49^. CIP is suggested as an alternative when first-line agents cannot be used, or as first choice when pyelonephritis is suspected, typically alongside a single parenteral dose of 3GC. Despite no antibiotic exposure in the year prior to enrollment, more than one-third of participants carried gut *E. coli* resistant to at least one of the three antibiotics. Thus, our data are in line with studies suggesting that carriage of resistant *E. coli* can occur independently of antibiotics use, though their acquisition may be associated with recent hospitalization, international travel, diet, etc.^50–52^

Interestingly, strains from pandemic MDR clonal groups ST131-*H30* and ST1193 were more often a predominant sub-population of gut *E. coli* than other resistant strains. This supports the hypothesis that the unusually broad global spread of these clonal groups is linked to their high fitness as gut colonizers^12,45,53–55^. Another striking finding of our study is that the carriage of ST131-*H30* and ST1193 is more common among older and younger women, respectfully. The biological reasons for these differences remain unclear. Considering that the subgroups’ resistance is similar, the key selective factor could be the host immunity status. Regardless, this variation has clinical significance. The 18-month UTI incidence among ST131-*H30* carriers was twice the overall rate (1 in 5 women), and in those aged 70+, more than one-third developed ST131-*H30*-associated UTIs within 18 months – the highest rate among all subgroups. Independently of resistance, ST131-*H30* is known being associated with complicated and severe UTIs, including recurrent cystitis, pyelonephritis, urosepsis, and infections in immunocompromised or diabetic patients^56^. In contrast, no such association was shown so far for ST1193, and its carriers had average UTI rates, likely to reflect differences in clonotype-specific urovirulence mechanisms.

Overall, our findings underscore the importance of predicting UTI risk not only by detecting gut *E. coli* but also by identifying specific clonal groups. Like many human pathogens, *E. coli* is clonal in nature, with most strains belonging to a limited set of genetically distinct groups, each differing in both virulence and resistance profiles^57,58^. While we did not examine in detail the fecal occurrence of other major clonotypes, our data highlight the value of sub-species clonal microbiome profiling – a “clonobiome” analysis approach^59^. We demonstrated that *E. coli* clonal diversity can be resolved directly in samples by shotgun metagenomic sequencing, which provides deeper resolution than high-throughput amplicon-based clonotyping^59^.

Beyond risk stratification, antibiotic susceptibility prediction is arguably even more critical once infection develops. In our study for the pathogen-defined UTI subgroup (N=134), within 18 months of fecal sampling, the resistance profiles of gut *E. coli* showed strong concordance with UTI pathogen susceptibility to TMP/STX, CIP, and 3GC, with excellent discrimination based on AUC values [49]. Using PPV in our cohort, a fecal *E. coli*-guided empiric (pre-culture) antibiotic choice would have resulted in drug-bug mismatch rates of only 1.9% for TMP/STX or CIP, and 0.8% for 3GC in predicted “susceptible” cases. In contrast, if these antibiotics had been prescribed empirically to all patients in our UTI cohort, mismatch rates would have been multifold-fold higher based on actual urine pathogen resistance – 14.2% for TMP/STX or CIP, and 3.7% for 3GC (see Table 1). Overall, fecal-based susceptibility prediction would be most impactful for TMP/STX and CIP, whose clinical use has declined at least in part due to increasing resistance. At the same time, the inappropriately increasing use of 3GC could be significantly limited by the fecal *E. coli* profiling. The impact of reducing drug-bug mismatches and the overuse of 3GC and other broader-spectrum antibiotics would be even more pronounced in populations and geographic regions with a substantially higher risk of MDR infections than the population in our study.

The correlation between gut and urinary *E. coli* resistance stems from the fact that UTI-causing strains arise from gut colonizers. While previous studies have demonstrated clonal concordance between fecal and urinary *E. coli* isolates, they vary widely in their estimates of gut colonization duration and have not leveraged baseline fecal profiles for prospective, long-term prediction of UTI risk and uropathogen susceptibility at a population scale^37,38,52,60–68^. For instance, Søraas et al.^52^ demonstrated that community-acquired ESBL carriage declines within six weeks of returning from travel, suggesting that gut *E. coli* populations are dynamic and influenced by environmental factors such as travel and diet – though clone-specific persistence, particularly for MDR ST131, may extend considerably longer. While multiple *E. coli* strains can co-colonize the gut simultaneously, the use of highly granular sub-MLST level of *fumC/fimH* (CH) clonotyping makes it unlikely that distinct strains sharing identical CH types were systematically misidentified as a single clone in our cohort. This is further supported by the consistent phylogenetic clustering of all 42 WGS-confirmed fecal-urinary pairs relative to an expanded set of unrelated strains of the same ST. Our study indicates that, depending on the relative abundance and profile of resistant strains, the probability of their continuous gut persistence for up to 18 months is at least 55-80%, based on the identity of the original fecal and follow-up UTI strains from the same individual. However, the study is limited by lack of longitudinal fecal sampling, limited precision in small subgroups and absence of analysis of exposure to antibiotics, dietary changes, travel or hospitalization for any reason during the follow-up. At the same time, because the gut typically harbors multiple *E. coli* strains, co-colonization may explain some apparent mismatches between fecal resistance profiles and UTI-causing strains. This is particularly relevant given that 55 of 177 samples with multiple antibiotic resistance contained a mixture of distinct resistant strains rather than a single MDR clone. In polyclonally colonized individuals, the fecal resistance profile reflects the combined repertoire of co-existing strains, only one of which may cause UTI, potentially affecting both concordance estimates and NPV in either direction. Alternatively, strain turnover may lead to replacement of the originally detected resistant strain and in fact, the lower UTI rates among the carriers of the low abundant strains could be due to a high loss rate among the latter from the gut. In a few cases, however, infections were likely caused by resistant strains absent from the initial fecal sample but acquired later. Since no significant association was detected between *E. coli* CH clonotype richness in ST131-*H*30-carrying fecal samples and UTI risk during follow-up (P=.254), these findings suggest that it is the presence of specific high-risk clones rather than overall clonal diversity per se that drives UTI susceptibility in this population. Future studies should clarify the gut colonization dynamics of uropathogenic *E. coli,* and further improvement in fecal test accuracy could be achieved by incorporating information on the abundance and urovirulence potential of different *E. coli* strains or clonal groups in the gut.

Plasmid loss or acquisition can also result in resistance discordance between fecal and urinary isolates independently of clonal dynamics. In our dataset, we identified one such case among 75 typed UTI episodes, in which the uropathogenic ST1193 strain had lost a blaCTX-M-55-carrying plasmid present in its fecal counterpart. In-depth plasmid analysis in larger discordant datasets would be valuable to further clarify the contribution of plasmid dynamics to resistance prediction accuracy.

Further work is needed to determine whether incorporating more features such as composition of other species in microbiome, more detailed strain diversity and abundance, virulence factors presence, and specific resistance gene profiles could improve UTI risk and treatment prediction. Validation studies should also test whether the fecal testing can extend to a broader antibiotic panel and to other host populations, including younger women, children, men, individuals with specific comorbidities as well as hospitalized patients, from geographically and socioeconomically diverse backgrounds. Interventional studies such as targeted decolonization trials of high-risk clonal groups would be needed to establish whether modifying the gut *E. coli* reservoir directly reduces UTI incidence and would provide the causal validation that the present observational study cannot. However, despite limitations, our findings support the further investigation of gut *E. coli* profiling for personalized UTI management. Moreover, they raise the prospect of preventive strategies – such as targeted decolonization of high-risk strains – that could reduce UTI burden and curb the community spread of MDR pathogens.

## Methods

### Study design and participants

This prospective cohort study from May 1, 2021 through December 31, 2021 was conducted among women aged 50 years and older enrolled in Kaiser Permanente Washington (KPWA). One-year membership prior to enrollment was required to clinical and demographic data prior to baseline. Besides enrollment, age and gender, exclusion criteria included any antibiotic prescription, UTI and/or placement in a long-term care facility within a year prior to enrollment in the study. The procedural details pertaining to the mailing of study kits to participants are described in the previous manuscript^25^ and in Supplemental Methods.

### Ethics approval and consent to participate

This study enrolled only adult participants; no minors were involved at any stage of recruitment, sampling, or analysis. The research protocol was reviewed and approved by the Kaiser Permanente Washington Research Institute Institutional Review Board (IRB), approval number **1563877**. All study procedures were conducted in accordance with the IRB-approved protocol, institutional ethical guidelines, and the principles outlined in the Declaration of Helsinki. Participants received a consent information sheet with the mailed self-collection kit. Voluntary return of the completed kit constituted implied consent to participate, as explicitly approved by the IRB. *Consent for publication* was not required since this study includes no identifiable individual participant data.

### Processing fecal samples and typing of fecal E. coli

was performed as described previously^25^ and is depicted in **Supplemental Figure S10** and in Supplemental Methods. Briefly, all fecal samples were plated on pre-poured HardyCHROM™ UTI agar plates (Hardy Diagnostic, USA) or plates prepared from HiChrome^TM^ UTI Agar (HiMedia Laboratories Pvt, Ltd.), both without any antibiotics and supplemented with ciprofloxacin (CIP, 0.5 mg/L),trimethoprim/sulfamethoxazole (TMP/SXT, 4/76 mg/L), ceftazidime (CAZ, 8 mg/L), cefotaxime (CTX, 2 mg/L) using standard quadrant plating technique ^69^. Plates were incubated for 16-20h at 37°C, and single colonies (SCs) morphologically identified as potential *E. coli* were cultured, saved and tested further for (a) resistance to CIP, TMP/SXT and 3^rd^ generation cephalosporins (3GC – ceftazidime, CAZ, and cefotaxime, CTX), (b) clonality based on sequencing of four loci – *fumC*, *fimH*, *gyrA* and *parC*, as described previously ^25^. Detailed description of sample processing and typing of *E. coli* can be found in **Supplemental Methods** and **Supplemental Table S4**.

### Processing clinical urine samples and typing of urine E. coli

If a study participant submitted a urine sample to KPWA clinical laboratory, a routine urinalysis test was performed, followed by culture and sensitivity testing if required. Starting February 2022, the UW laboratory was provided an aliquot of the urine sample, which was processed using the same protocol as fecal sample described above, adding *E. coli* clonotype identity where applicable to sample information. The time lapse between the beginning of the study (May 2021) and the start of availability of clinical urine samples to the UW lab accounts for the lack of clonal information for some *E. coli*-caused UTIs.

### UTI identification

The incidence of urinary tract infections in study participants was deduced from their electronic health records (EHRs). Records were examined once all participants had surpassed 18 months following the submission of the fecal sample. UTI was characterized based on the combinatorial criteria outlined in Bruxvoort et al^7^, including UTI diagnosis codes, antibiotic prescriptions, urinalysis test results, and culture findings. Urine culture and susceptibility data were available for 134 of 184 UTI cases; the remaining 50 cases had no susceptibility data recorded, either due to absence of urine culture submission or negative/contaminated culture results. We did not identify obvious demographic or clinical differences between cases with and without culture data; however, a formal comparison was limited by the data available to us, comprising diagnosis, urinalysis, culture and susceptibility results, medications, and age.

### Search for urinary E. coli clonotypes within fecal samples

Fecal samples were examined to identify the presence of the *E. coli* clonotype potentially responsible for UTI in the participant. To begin with, if the clinical urine *E. coli* was resistant to CIP, TMP/SXT, and/or 3GC antibiotics, and the matching fecal sample contained *E. coli* with matching resistance profile, the clonal profile was compared too if the urinary *E. coli* was available for clonal typing.

In case the urinary *E. coli* was CIP-, TMP/SXT-, 3GC-sensitive, we used following approach to try to identify its presence among fecal *E. coli*. Firstly, the uropathogenic *E. coli* strains isolated from clinical urine samples were subjected to testing for resistance against a broader spectrum of antibiotics, including tetracycline (16 mg/L), cefazoline (4 and 8 mg/L), chloramphenicol (16 and 32 mg/L), kanamycin (12.5 and 25 mg/L), gentamicin (15 mg/L), spectinomycin (15 mg/L), streptomycin (50 mg/L), trimethoprim (8 and 16 mg/L), and sulfamethoxazole (80 and 160 mg/L). Subsequently, fecal samples were plated on the antibiotic of interest to search for isolates exhibiting the same resistance profile.

If urinary *E. coli* did not have unique resistance profile, the fecal sample was plated on UTI agar to isolate and type potential *E. coli*. Overall, up to 50 single *E. coli* colonies were screened per sample, with minimum 33 colonies for samples where *E. coli* of interest was not found using this approach.

The fecal samples where *E. coli* of interest were not found were subjected to final screening using qPCR approach. Fecal samples were plated on McConkey agar to obtain at least 1000 single colonies. DNA was isolated from pooled colonies, and clone-specific qPCRs was performed to detect the presence of clone of interest in a pooled sample. If the SNPs’ presence was detected, individual colonies were checked further as described above.

### UTI and fecal E. coli isolates comparison using whole genome ***sequencing*.**

Fecal and urine *E. coli* isolates from the same participant with matching *fumC-fimH* (CH) type had their genomes sequenced on an Illumina MiSeq platform using the MiSeq 600 cycle v3 kit, following the manufacturer’s guidelines. Genomic DNA libraries were prepared with the Nextera XT Library Prep Kit (Illumina, CA). Raw data were uploaded to the Enterobase database (https://enterobase.warwick.ac.uk/) for genome assembly, allele calling, and wgMLST and cgMLST assignments (Enterobase uberstrain names are listed in Supplemental Data). To investigate the phylogenetic relationships among isolates within the same clonotypes, we analyzed the sequences of cgMLST loci that varied between them. Potential polymorphisms were manually confirmed by examining raw reads and performing SNP-specific qPCR if necessary. For each fecal-urinary pair, a separate SNP-based phylogenetic tree was constructed using MEGA12, incorporating up to 14 of the closest publicly available genomes of the same ST retrieved from EnteroBase; full details are provided in the Supplemental Methods.

### Metagenomics analysis

A subset of 36 pairs of fecal and clinical urine samples was analyzed to assess the relative abundance of *E. coli* among enterobacteria, as well as the presence and prevalence of specific *E. coli* clones. The detailed procedure is described in the Supplemental Methods. Briefly, samples were plated on MacConkey agar to obtain 10³-10⁴ colonies per plate. DNA extracted from pooled colonies was used for shotgun metagenomic sequencing, following the same procedure described above for individual *E. coli* whole-genome sequencing. The raw reads were analyzed for species composition, urinary *E. coli* clone abundance and *E. coli* clonal composition using an in-house pipeline described in Supplemental Methods.

### Statistical analysis

Chi-square tests and the cci command were used for pairwise 2×2 comparisons, and logistic regression was used to analyse the prevalence of different *E. coli* clones and UTI incidence across age groups.

Continuous variables (e.g., *E. coli* abundance) were compared across categories using the Student’s t-test. Diagnostic accuracy metrics (sensitivity, specificity, PPV, and NPV) were calculated using the *diagt* command, and AUC values were estimated using *roctab*. All analyses were conducted in Stata 14.2 (StataCorp, College Station, TX, USA). Subgroup analyses were exploratory and were not corrected for multiple comparisons; findings should be interpreted accordingly.

## Authors’ contributions

**E.V.S.** and **J.D.R.** designed the study, secured funding and supervised the project. **E.V.S.** wrote the original draft. **V.T.** managed the project at UW, performed data analysis and contributed to study design and writing the original draft. **D.C.** performed experimental work, metagenomics and sequencing analysis and assisted with data interpretation. **L.L., I.B., Y.S., T.C.B., T.S., J.H., I.P.** and **S.P.** carried out wet-lab experiments and contributed to data generation. **V.B.** conducted electronic medical record (EMR) data analysis. **J.S.** performed data analysis and generated violin plot visualizations. **G.S.D.** and **S.Y.T.** contributed to data analysis and provided critical revisions to the manuscript. **L.T.M.** provided programming support and data infrastructure development for KPWA. **E.H.** served as KPWA project manager and coordinated administrative and operational aspects of the study. All authors reviewed and approved the final manuscript.

## Data Availability

All relevant data are within the manuscript and its Supporting Information files. The whole genome sequencing data are available in the Enterobase database (https://enterobase.warwick.ac.uk/). The shotgun metagenomics data are in NCBI. All submission IDs are provided in Supplemental Data file.

## Acknowledgments

We would like to express our gratitude to Drs. James Johnson, Steve Moseley and Lance Price for critically reading the manuscript and providing important suggestions that improved its content and conceptual vision.

## Supplemental Information Inventory

1. **Supplemental Figure S1. Analysis of gut *E. coli* carriage and UTI incidence by age.** Number of participants, carriage rates and UTI incidence rates were aggregated for every two-year age bracket, and the frequency was calculated as a percentage of the total number of participants within each two-year age range. For participants 90-98 years of age numbers were aggregated into one bin. Plotted frequencies were used to calculate either the best-fit trend (grey dotted line), or linear trend (orange line), with R^2^ value shown on the graph in grey and orange, respectively. (A) Overall age distribution among study participants. (B) Prevalence of fecal samples with *E. coli*. (C) Prevalence of fecal samples with *E. coli* resistant to CIP. (D) Prevalence of fecal samples with *E. coli* resistant to TS. (E) Prevalence of fecal samples with *E. coli* resistant to 3GC. (F) Prevalence of fecal samples with *E. coli* from ST131-H30. (G) Prevalence of fecal samples with *E. coli* from ST1193.
2. **Supplemental Figure S2. Venn diagram showing the distribution of CIP-R, TMP/STX-R, and 3GC-R *E. coli* in fecal samples**. Overlaps indicate the presence of *E. coli* resistant to multiple antibiotics within the same fecal sample, regardless of whether the resistances are carried by the same or different clones.
3. **Supplemental Figure S3. Comparative analysis of relative abundance of *E. coli* in fecal sample versus other bacteria.** (A) Various patterns of relative species abundance based on the four-quadrant streaking of the fecal samples on UTI agar. *E. coli* produce pink colonies. (B) Correlation between *E. coli* abundance in feca samples estimated by growth on chromogenic UTI agar and determined by 16S sequencing. The size of a bubble reflects the number of samples. A linear trendline was plotted for all samples except one marked in color (*E. marmotae)*, with R^2^ value indicated on the graph. (C) Relative abundance of *E. coli* and other bacterial species in fecal samples by 16S. The X-axis represents individual fecal samples, while the Y-axis shows the relative abundance of bacterial species as a percentage of the total bacterial composition in each sample.
4. **Supplemental Figure S4. Distribution of different *E. coli* abundancies in fecal samples.** (A-B) Combined violin/box plot distribution of *E. coli* abundances in fecal sample relatively to Gram-negative (A) or total (B) bacterial grown on UTI agar, split by the different age groups. Sample sizes for each age group are shown in parentheses. Mean abundances (Mean ± SEM %) are displayed above the plots, with no significant difference between the groups. (C-D) Distribution of CIP-R *E. coli* abundance compared to average *E. coli* abundance. (C) All CIP-R growth level relative to any *E. coli* growth is plotted on primary vertical axis, whereas average *E. coli* growth relative to Gram-negative (G-) or overall (Total) bacterial growth in these subsets of samples is plotted on a secondary vertical axis, with trendlines and R^2^ values representing linear fit of the data. Error bars represent standard error. (D) Same as (C) for different CIPREc clonal groups, with average *E. coli* growth relative to Gram-negative growth only.
5. **Figure S5. UTI incidence rates and uropathogens’ characteristics.** Plotted here are overlaying frequencies as follows: total number of UTIs occurring within 1-18 months from submitting fecal samples in grey; positive urine culture in dark-grey; *E. coli* in urine green; *E. coli* resistant to FQ, TS and/or 3GC in orange; *E. coli* resistant to FQ, TS and/or 3GC and not found in fecal sample in red.
6. **Supplemental Figure S6. Analysis of core-genome sequences for paired fecal (F) and corresponding clinical urine (CU) isolates.** Molecular phylogenetic analyses were performed using the Maximum Likelihood method implemented in MEGA12, generating SNP-based phylogenies for *E. coli* fecal-urinary pairs. For each pair, a separate phylogenetic tree was constructed incorporating the closest publicly available genomes of the same sequence type retrieved from EnteroBase.
7. Supplemental Figure S7. Violin/box plot of abundancies of different resistant *E. coli* vs. total *E. coli* in fecal samples of women with and without UTI. A) Among CIP-R *E. coli* carriers; B) Among TMP/SXT-R *E. coli* carriers; C) Among 3GC-R *E. coli* carriers; D) Among carriers of different clonal groups of CIP-R *E. coli*. Sample size is shown in parenthesis. Mean abundance (Mean ± SEM%) is displayed above the plots. *P* values for statistically significant differences are indicated above the solid lines.
8. **Supplemental Figure S8. Correlation between prevalence of clones identified by growth analysis and Metagenomics analysis.** For 36 fecal samples shotgun metagenomics was performed for pooled 1,000-10,000 single colonies grown on McConkey agar. UTI *E. coli* clone’s abundance among all fecal *E. coli* within each sample was determined both from metagenomic analysis and culture data, and compared on the graph, with the bubble size reflecting number of samples. Light grey bubble indicates cases where urinary *E. coli* was not found either by metagenomic analysis or by culture. Trendline and R^2^ value indicated the linear fit.
9. Supplemental Figure S9. Effect of abundance of resistant *E. coli* within fecal sample on prediction of uropathogen’s antibiotic susceptibility. PPV (prediction of susceptibility to antibiotic), NPV (prediction of resistance to antibiotic) and AUC (overall test performance) was calculated for different cutoff of resistant *E. coli* prevalence within fecal samples (aka, abundance).
10. **Supplemental Figure S10.** Flowchart outlining the workflow for processing baseline fecal samples and subsequent clinical urine isolates, including sample collection, culturing, sequencing, and downstream genomic analyses.
11. Supplemental Methods.
12. **Supplemental Table S1.** Clonotype distribution of multidrug-resistant CIP-R *E. coli* isolated from fecal samples. ST, sequence type, CC, clonal complex, H, *fimH* allele, Q, number of QRDR mutations in *gyrA* and/or *parC* known to confer CIP-resistance; for ST69-H27(2Q) and ST38-H5(1Q) and ST38-H65(3Q) *gyrA* and *parC* allele numbers are listed after comma to indicate different CIP-R clones.
13. **Supplemental Table S2**. UTI *E. coli* resistant to antibiotics.
14. **Supplemental Table S3**. Non-*E. coli* UTI uropathogens.
15. **Supplemental Table S4**. Primers used for CH typing, *gyrA*-*parC* sequencing.
16. **Supplemental Table S5.** Classification of participants with diagnosed UTI by uropathogen and baseline fecal *E. coli* carriage profile.
17. **Supplemental Data.** File containing participant-level data, including age, baseline fecal sample analyses, UTI occurrence, time elapsed since fecal sample submission (in months), urine sample characteristics (when available), and whole-genome sequencing and metagenomics data with corresponding NCBI and EnteroBase database entries.
18. **Supplemental R script.** In-house R script for identification of publicly available genomes most closely related to a query isolate based on core-genome SNP distances.

