## Supplemental Materials for "Personalized prediction of UTI risk and antibiotic susceptibility based on gut *E. coli*"

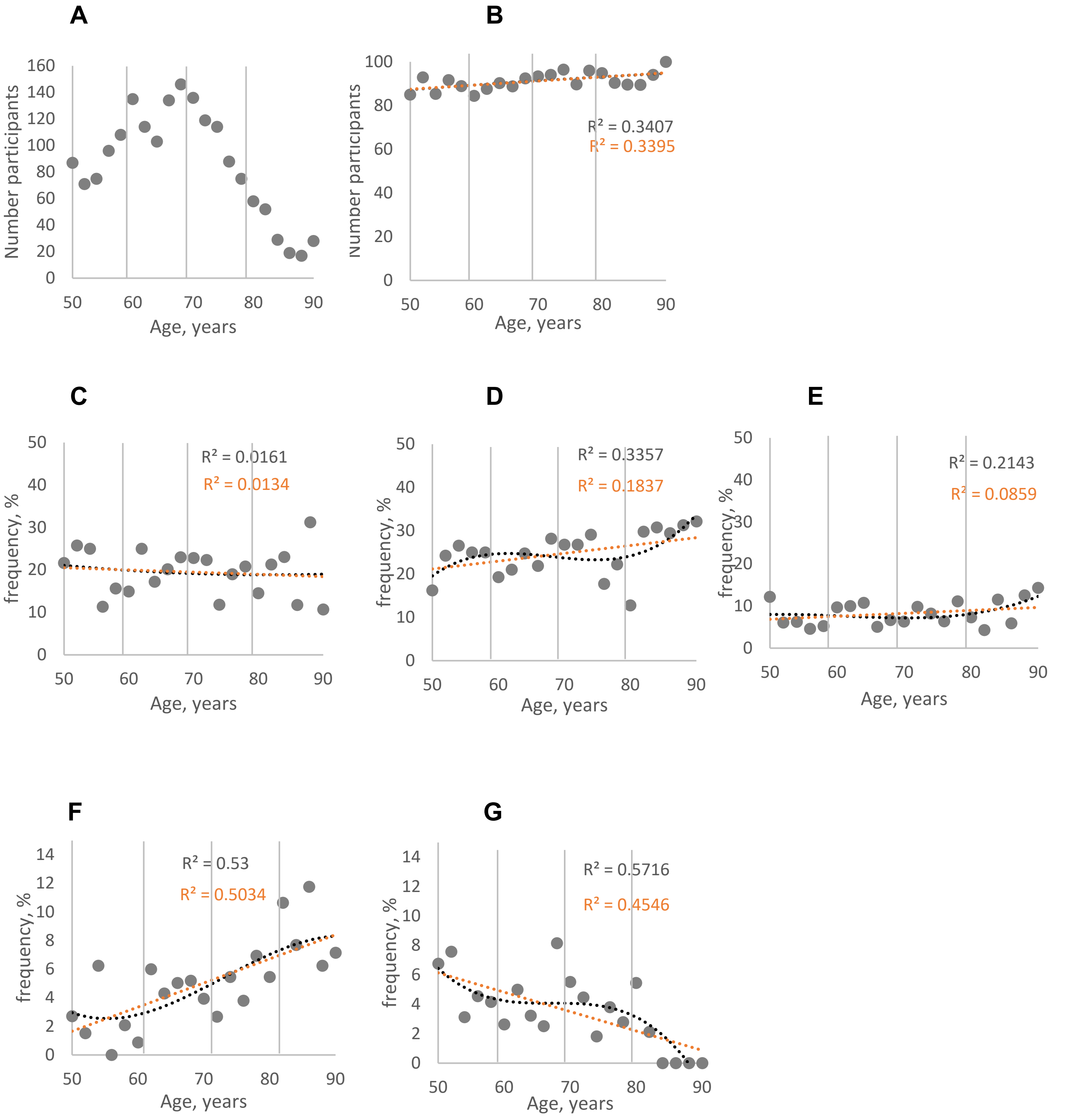


**Figure S1. Analysis of gut *E. coli* carriage and UTI incidence by age.** Number of participants, carriage rates and UTI incidence rates were aggregated for every two-year age bracket, and the frequency was calculated as a percentage of the total number of participants within each two-year age range. For participants 90-98 years of age numbers were aggregated into one bin. Plotted frequencies were used to calculate either the best-fit trend (grey dotted line), or linear trend (orange line), with $R^2$ value shown on the graph in grey and orange, respectively. (A) Overall age distribution among study participants. (B) Prevalence of fecal samples with E. coli. (C) Prevalence of fecal samples with *E. coli* resistant to CIP. (D) Prevalence of fecal samples with *E. coli* resistant to TS. (E) Prevalence of fecal samples with *E. coli* resistant to 3GC. (F) Prevalence of fecal samples with *E. coli* from ST131-H30. (G) Prevalence of fecal samples with *E. coli* from ST1193.

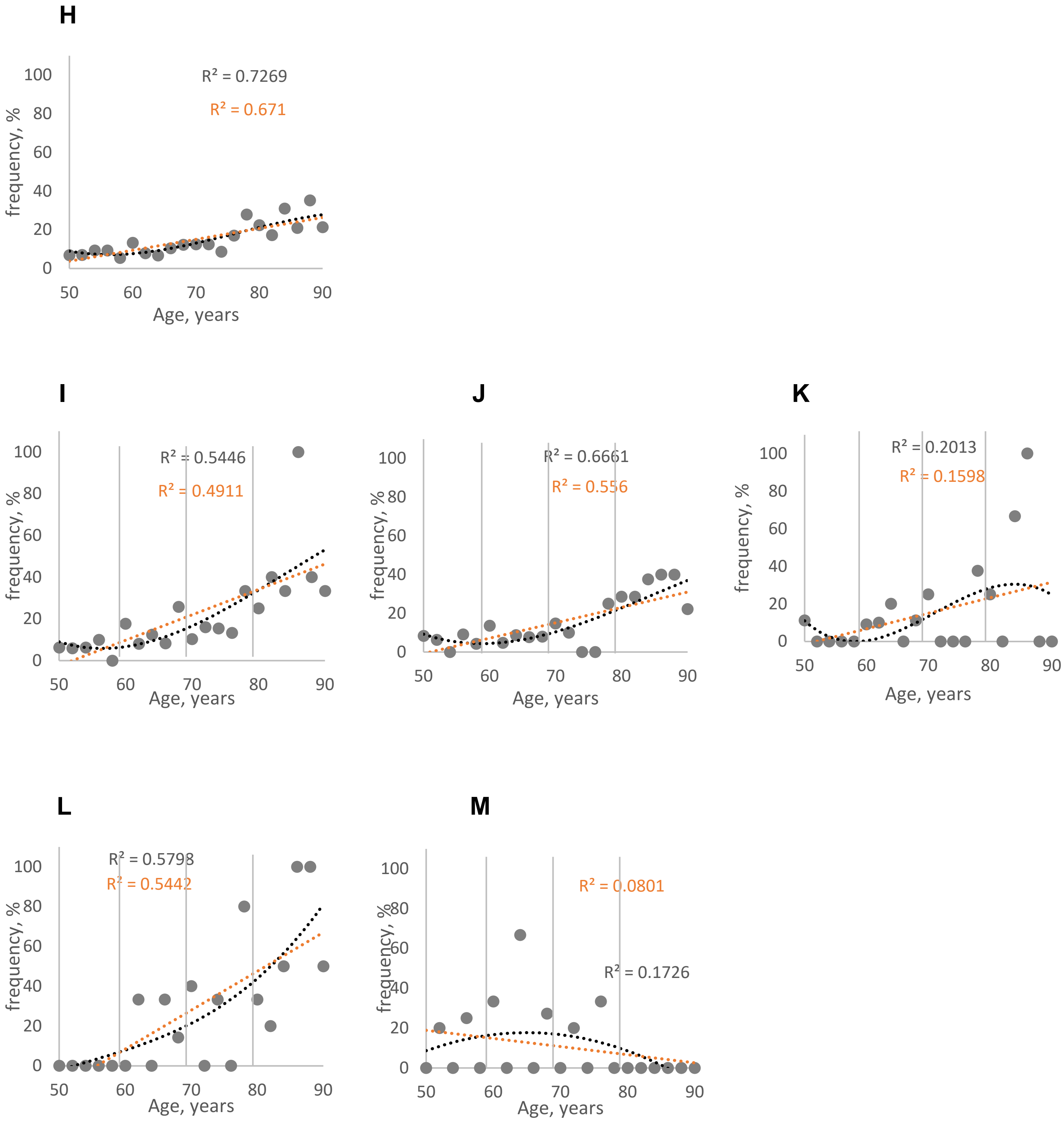


**Supplemental Figure S1. Analysis of gut *E. coli* carriage and UTI incidence by age22 (continued).** (H) Overall UTI incidence rate distribution by age. UTI incidence rates in resistant *E. coli* carriers: FQREC carriers (I), TSREC carriers (J), 3GCREC carriers (K), H30 carriers (L) and ST1193 carriers (M).

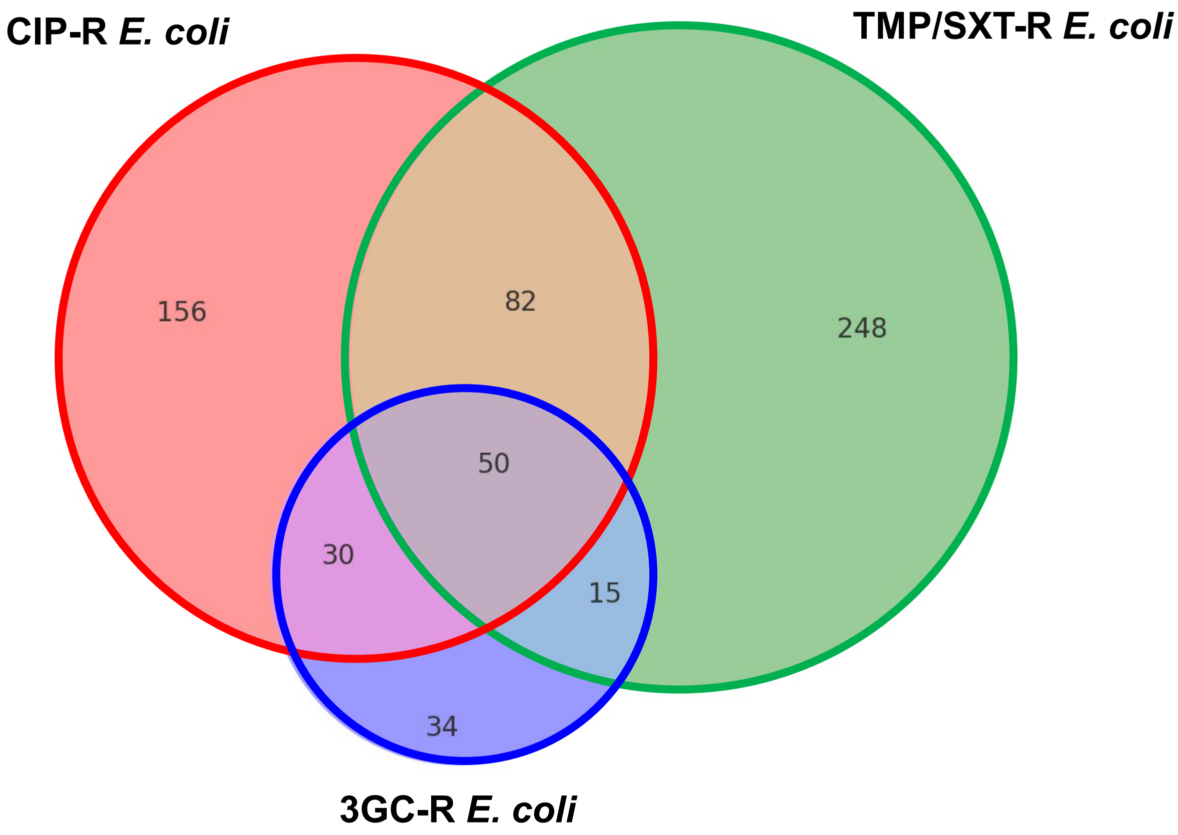


**Figure S2. Venn diagram showing the distribution of CIP-R, TMP/STX-R, and 3GC-R *E. coli* in fecal samples.** Overlaps indicate the presence of *E. coli* resistant to multiple antibiotics within the same fecal sample, regardless of whether the resistances are carried by the same or different clones.

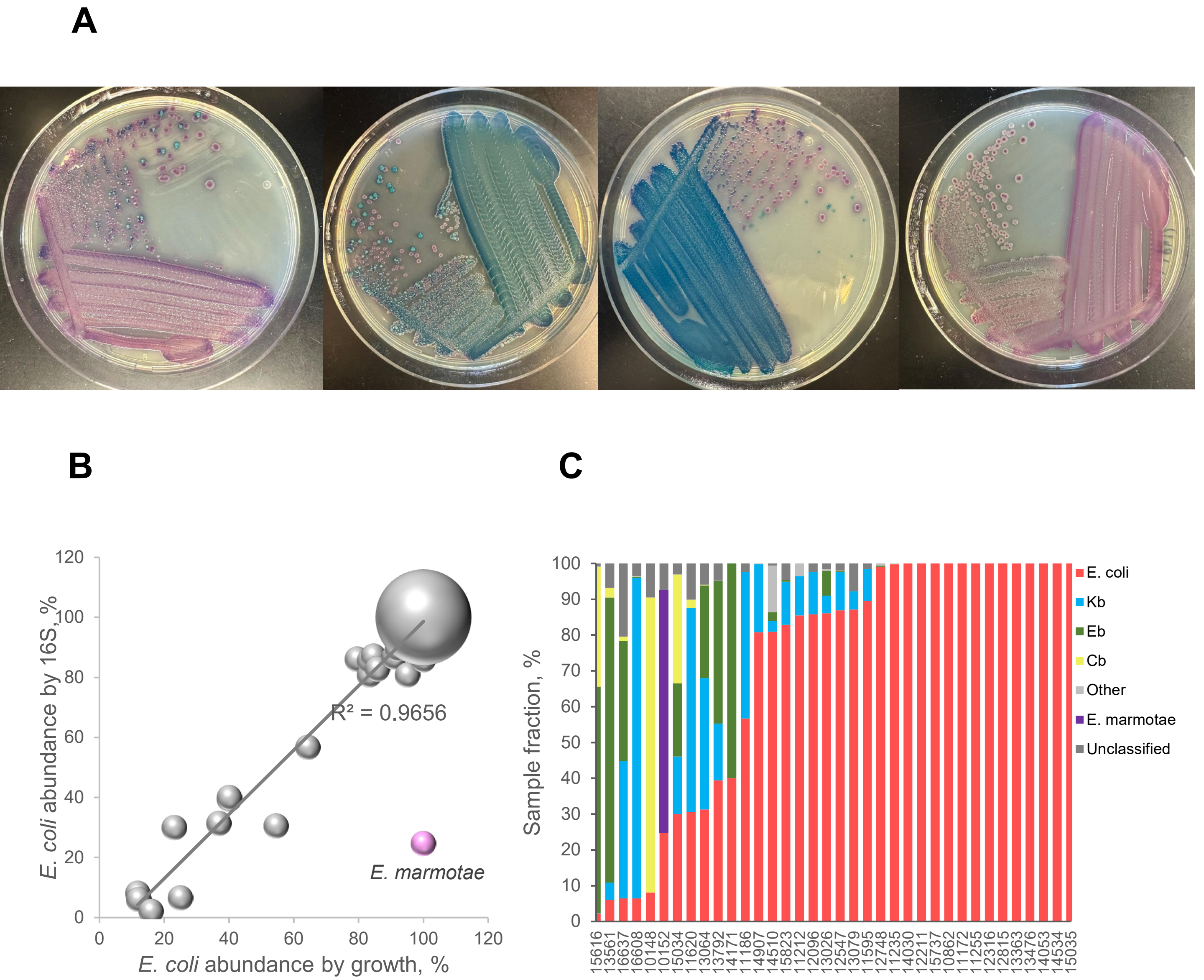


**Supplemental Figure S3. Comparative analysis of relative abundance of E. coli in fecal sample versus other bacteria.** (A) Various patterns of relative species abundance based on the four-quadrant streaking of the fecal samples on UTI agar. *E. coli* produce pink colonies. (B) Correlation between *E. coli* abundance in fecal samples estimated by growth on chromogenic UTI agar and determined by 16S sequencing. The size of a bubble reflects the number of samples. A linear trendline was plotted for all samples except one marked in color (*E. marmotae)*, with $R^2$ value indicated on the graph. (C) Relative abundance of *E. coli* and other bacterial species in fecal samples by 16S. The X-axis represents individual fecal samples, while the Y-axis shows the relative abundance of bacterial species as a percentage of the total bacterial composition in each sample.

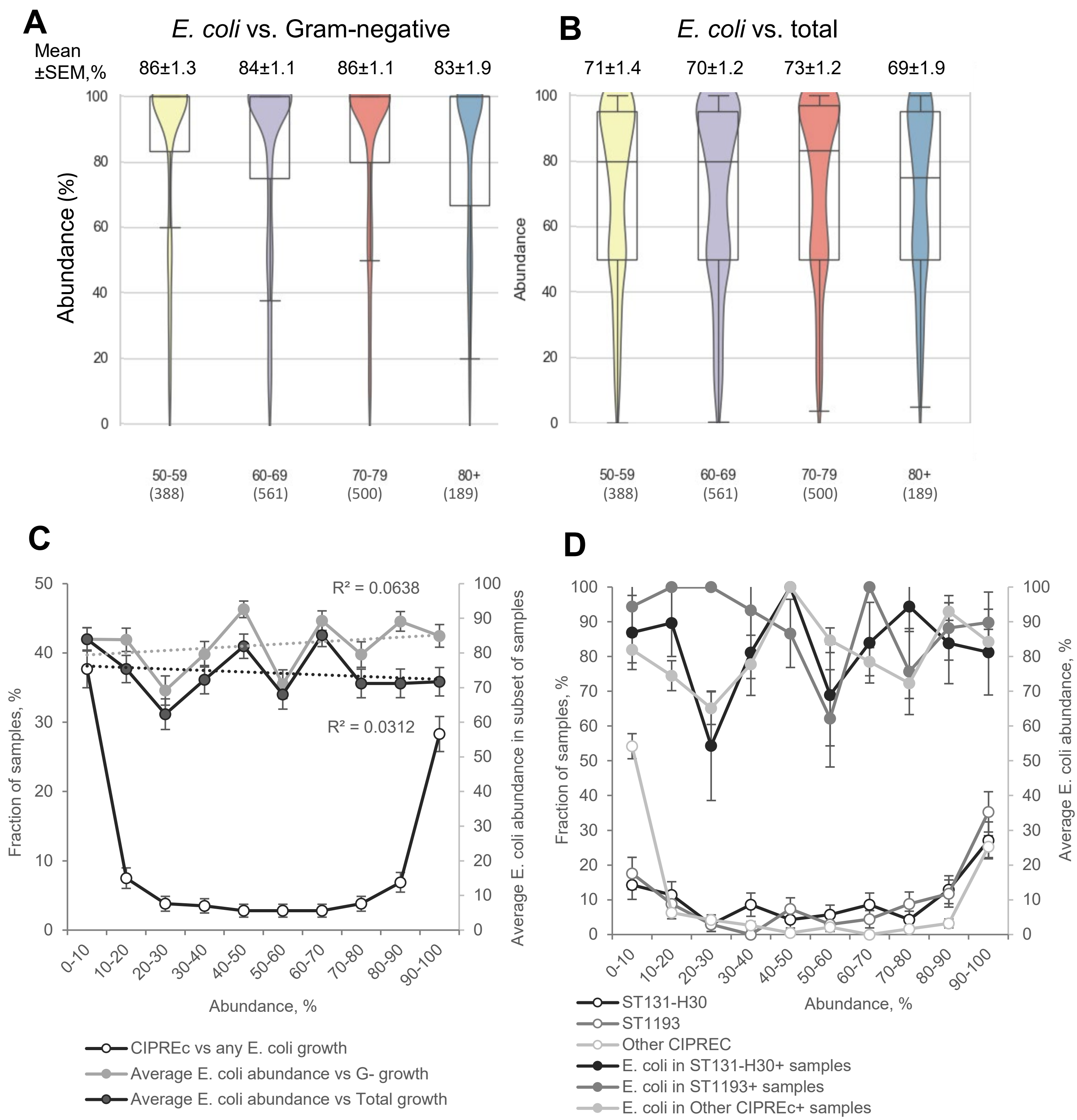


**Supplemental Figure S4. Distribution of different *E. coli* abundancies in fecal samples.** (A-B) Combined violin/box plot distribution of *E. coli* abundances in fecal sample relatively to Gram-negative (A) or total (B) bacterial grown on UTI agar, split by the different age groups. Sample sizes for each age group are shown in parentheses. Mean abundances (Mean ± SEM %) are displayed above the plots, with no significant difference between the groups. (C-D) Distribution of CIP-R *E. coli* abundance compared to average *E. coli* abundance. (C) All CIP-R growth level relative to any *E. coli* growth is plotted on primary vertical axis, whereas average *E. coli* growth relative to Gram-negative (G-) or overall (Total) bacterial growth in these subsets of samples is plotted on a secondary vertical axis, with trendlines and $R^2$ values representing linear fit of the data. Error bars represent standard error. (D) Same as (C) for different CIPREc clonal groups, with average *E. coli* growth relative to Gram-negative growth only.

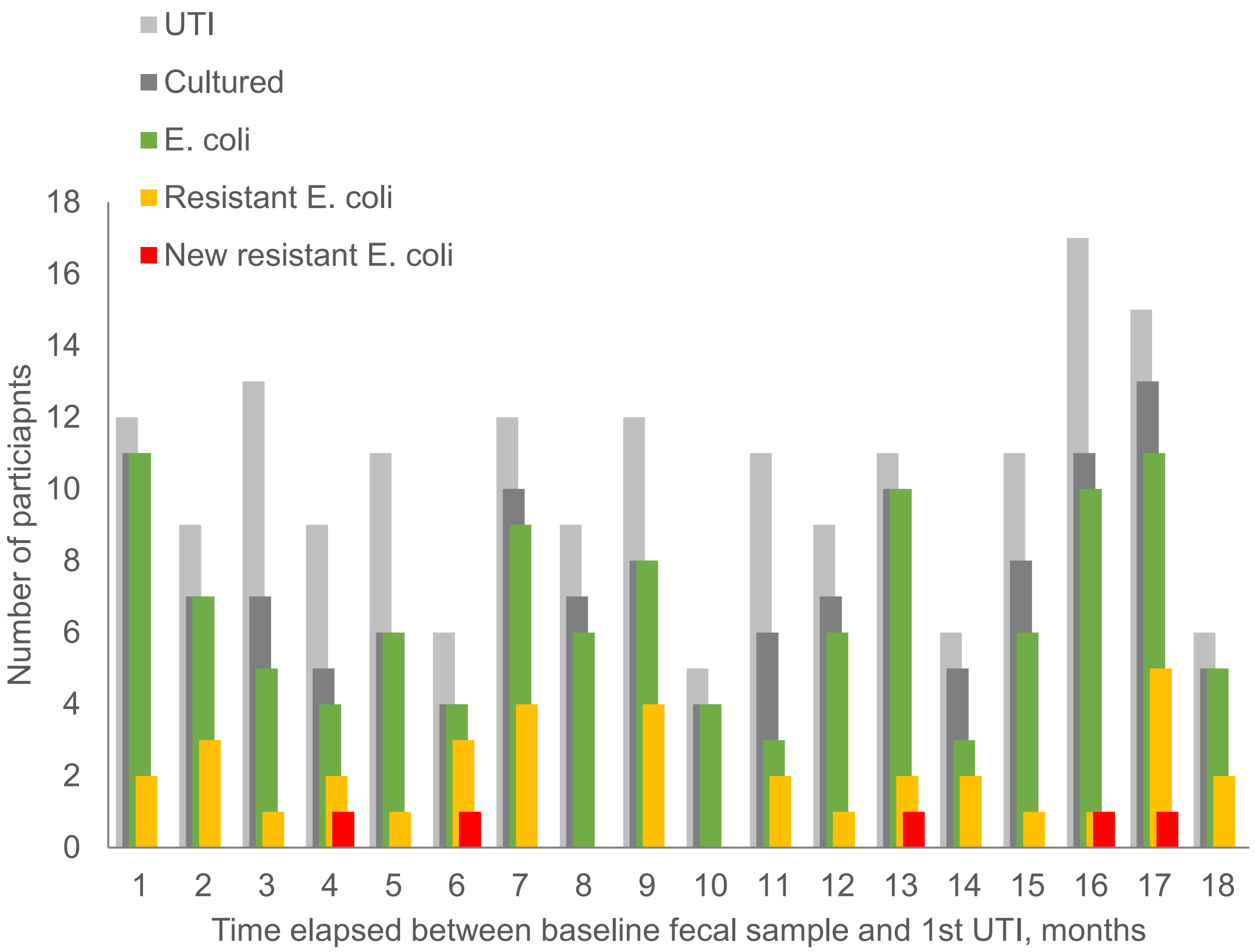


**Figure S5. UTI incidence rates and uropathogens' characteristics**. Plotted here are overlaying frequencies as follows: total number of UTIs occurring within 1-18 months from submitting fecal samples in grey; positive urine culture in dark-grey; *E. coli* in urine green; *E. coli* resistant to FQ, TS and/or 3GC in orange; *E. coli* resistant to FQ, TS and/or 3GC and not found in fecal sample in red.

**Supplemental Figure S6. Analysis of core-genome sequences for paired fecal (F) and corresponding clinical urine (CU) isolates.** Molecular phylogenetic analyses were performed using the Maximum Likelihood method implemented in MEGA12, generating SNP-based phylogenies for E. coli fecal-urinary pairs. For each pair, a separate phylogenetic tree was constructed incorporating the closest publicly available genomes of the same sequence type retrieved from EnteroBase.

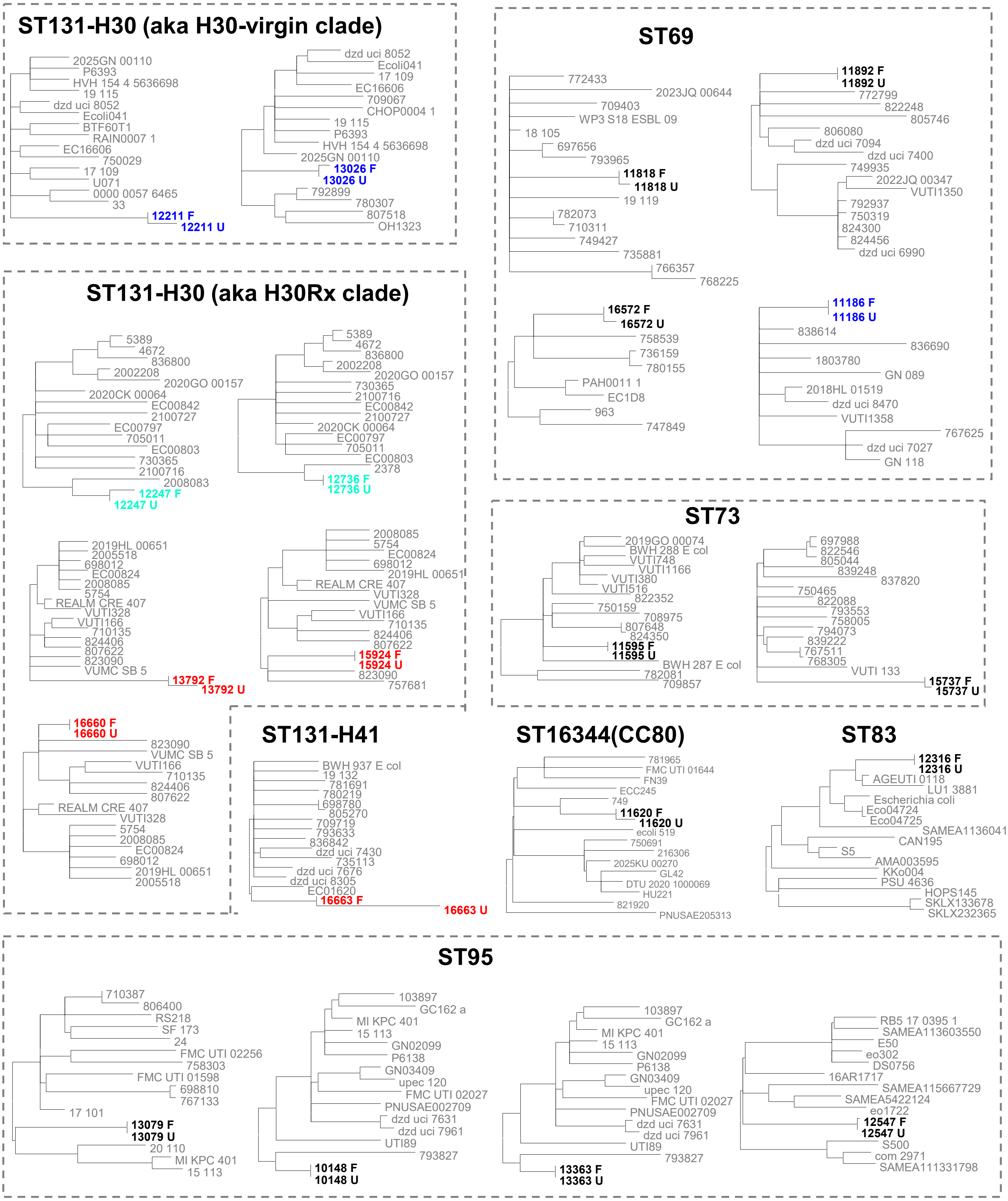

**Supplemental Figure S6 B.**

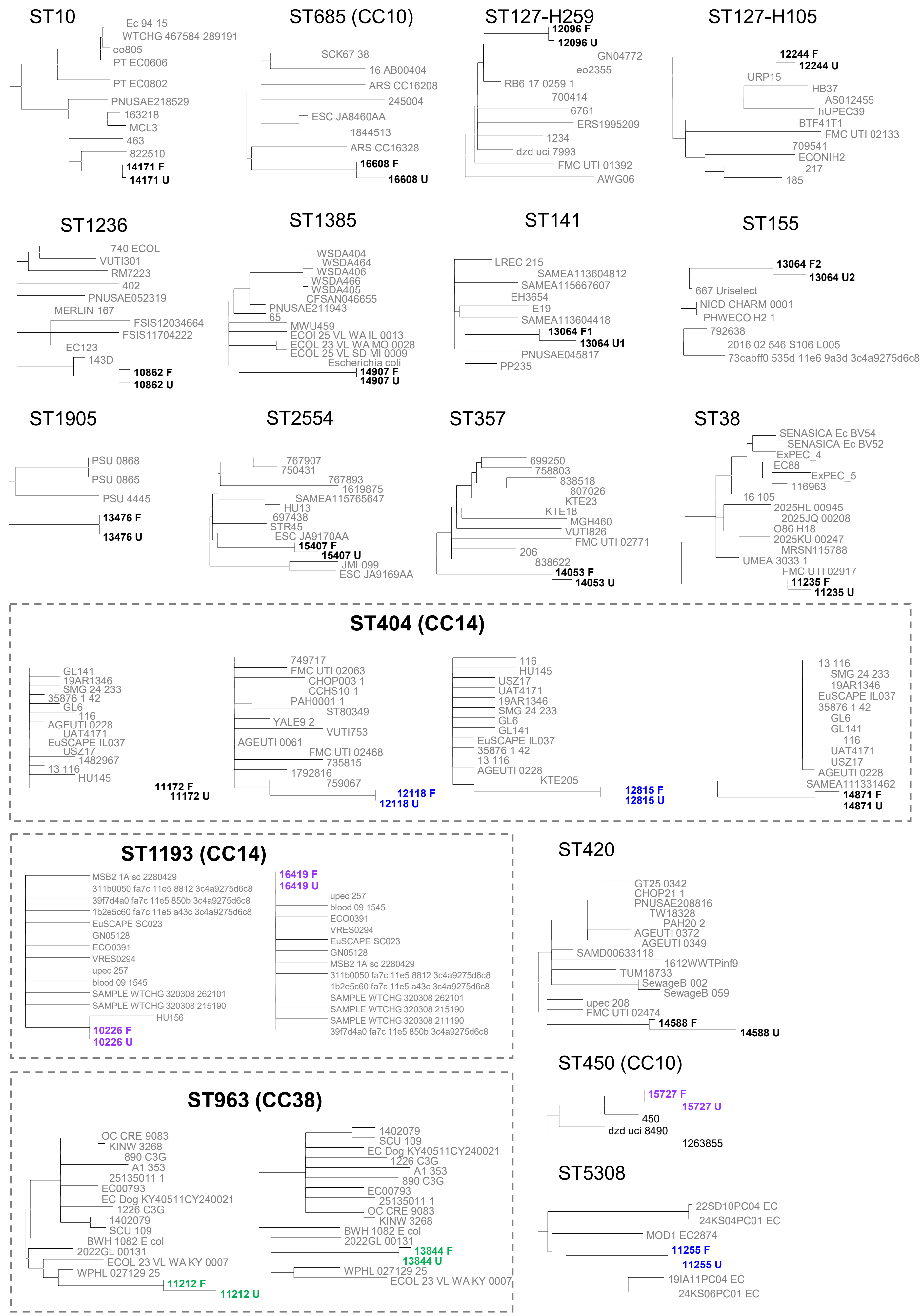

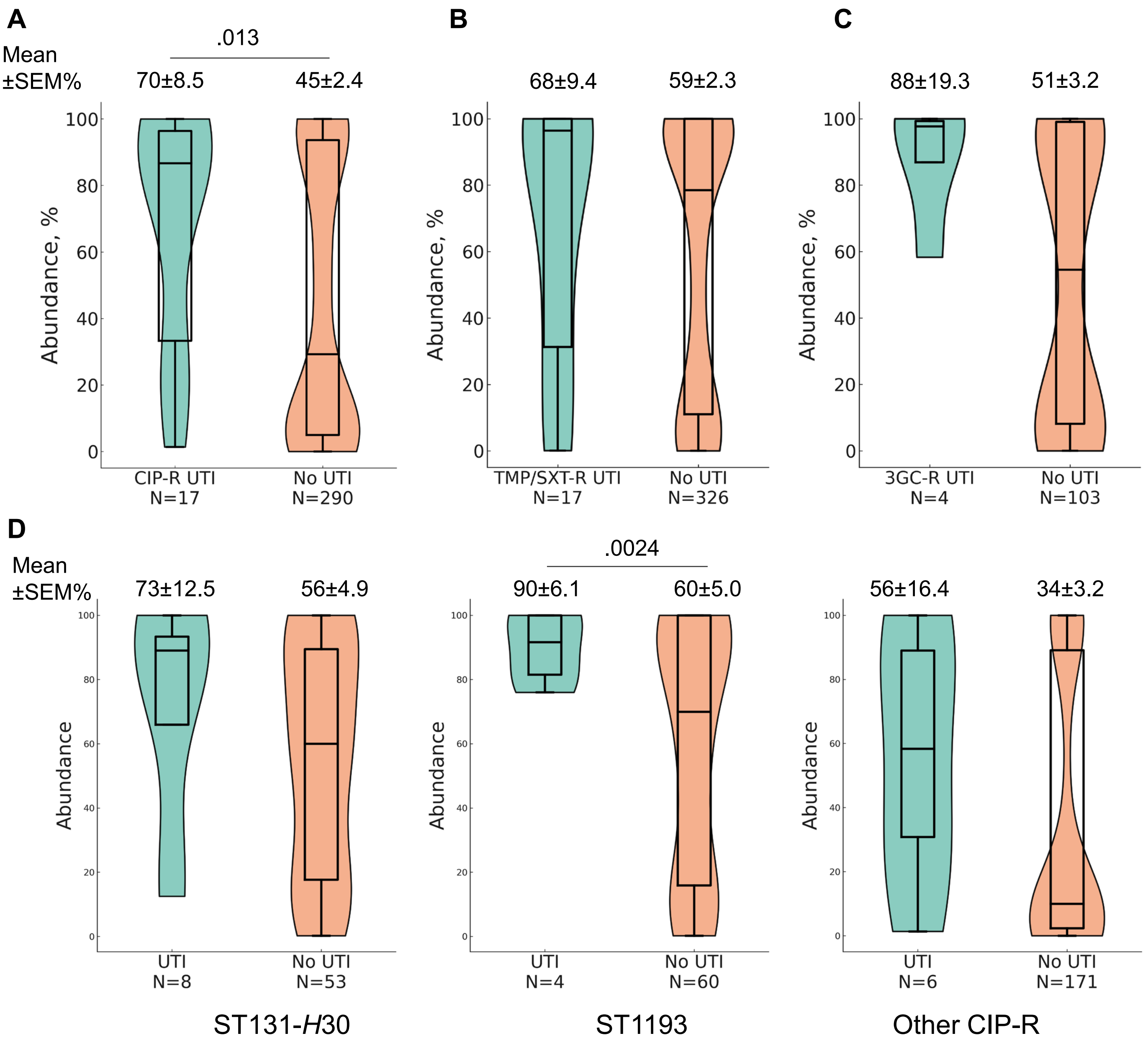


**Supplemental Figure S7. Violin/box plot of abundancies of different resistant *E. coli* vs. total *E. coli* in fecal samples of women with and without UTI.** A) Among CIP-R *E.* coli carriers; B) Among TMP/SXT-R *E.* coli carriers; C) Among 3GC-R *E.* coli carriers; D) Among carriers of different clonal groups of CIP-R *E. coli*. Sample size is shown in parenthesis. Mean abundance (Mean ± SEM%) is displayed above the plots. *P* values for statistically significant differences are indicated above the solid lines.

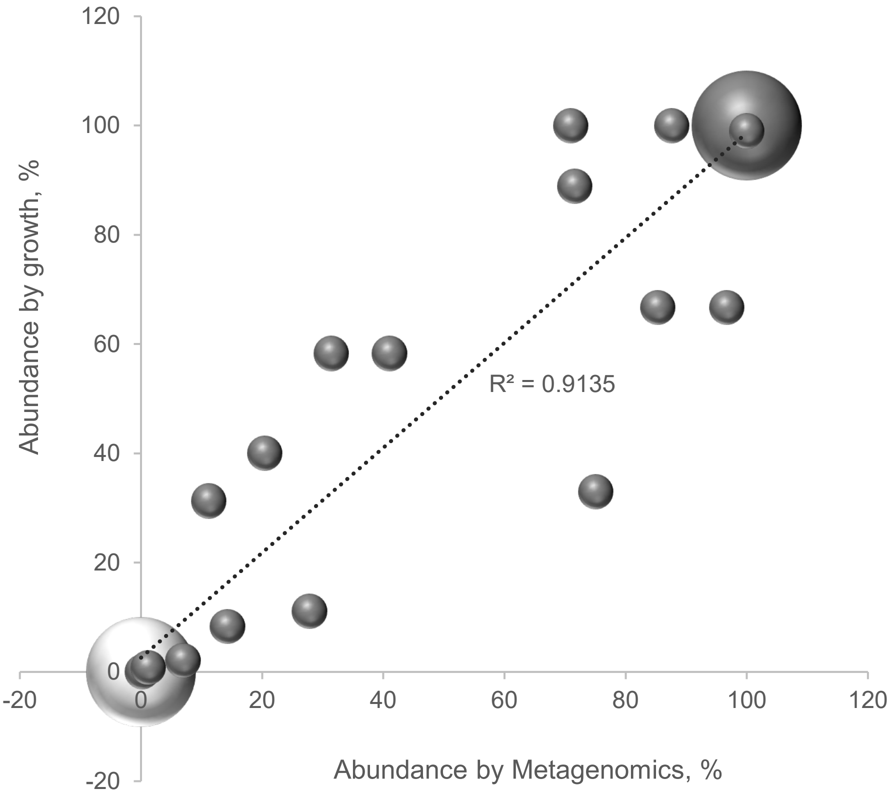


**Figure S8. Correlation between prevalence of clones identified by growth analysis and Metagenomics analysis.** For 36 fecal samples shotgun metagenomics was performed for pooled 1,000-10,000 single colonies grown on McConkey agar. UTI *E. coli* clone's abundance among all fecal *E. coli* within each sample was determined both from metagenomic analysis and culture data, and compared on the graph, with the bubble size reflecting number of samples. Light grey bubble indicates cases where urinary *E. coli* was not found either by metagenomic analysis or by culture. Trendline and $R^2$ value indicated the linear fit.

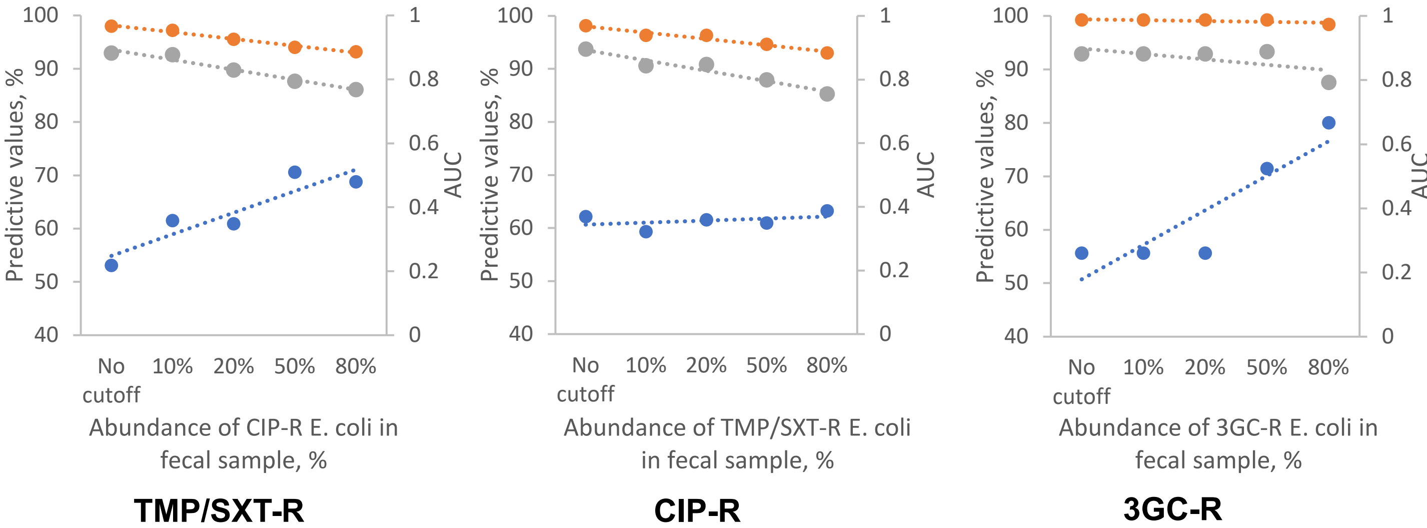


**Supplemental Figure S9. Effect of resistant *E. coli* abundance within fecal sample on prediction of uropathogen's antibiotic susceptibility**. PPV (prediction of susceptibility to antibiotic, orange circles), NPV (prediction of resistance to antibiotic, blue circles) and AUC (overall test performance, area-under-curve, grey circles) was calculated across different cutoffs of resistant *E. coli* prevalence within fecal samples (i.e., abundance). Dotted lines represent linear trendlines.

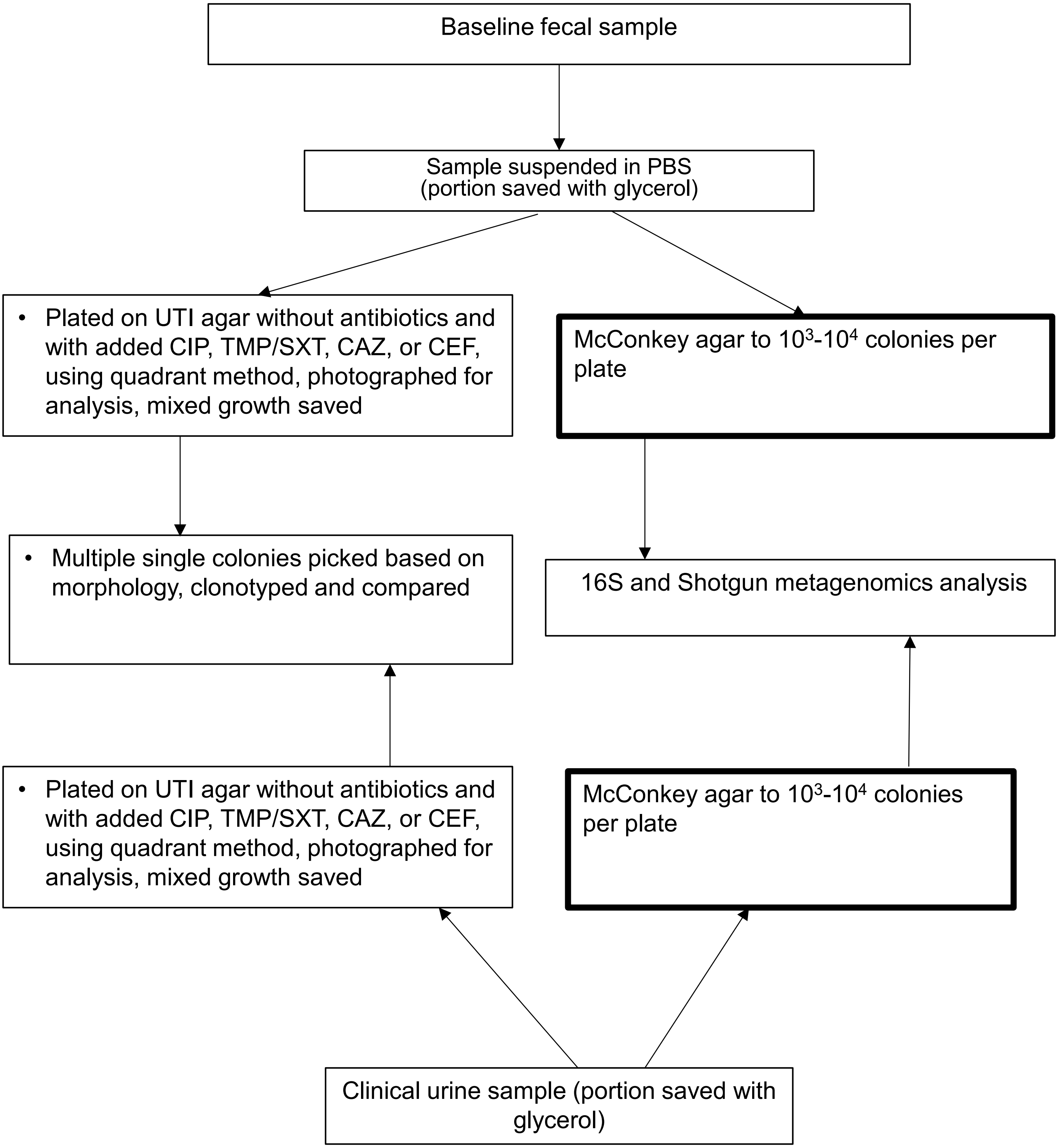


**Supplemental Figure S10.** Flowchart outlining the workflow for processing baseline fecal samples and subsequent clinical urine isolates, including sample collection, culturing, sequencing, and downstream genomic analyses.

### Supplemental Methods

#### Fecal sample collection

Fecal samples were collected via self-collection kits. The kits mailed to potential enrollees included a culture swab and tube, biohazard bag with absorbent material inside, a piece of bubble padding, a shipping box, invitation letter, consent information sheet and detailed instructions. Instructions specified the importance of labeling the collected sample with the date of collection and mailing the sample as soon as possible. Samples were received by KPRWHI survey team and stored at 4°C until picked up by UW processing team. For all samples the time lapse between collection date and processing date was on average 5.3±2.0 days, with 90% samples processed within a week from recorded sample collection.

#### Sample processing

The flow of sample processing steps is illustrated in Supplemental Figure S10. Fecal samples were self-collected using the FecalSwab™ Sample Collection and Preservation System for Enteric Bacteria by Copan Diagnostic Inc. (Carlsbad, CA, USA). At the processing start samples were visually assessed for the quality of fecal matter before being plated on four types of agar as described in our previous manuscript[1]. Pre-poured HardyCHROM™ UTI agar plates (Hardy Diagnostic, USA) were used for non-antibiotic plating of *E. coli*. The proprietary composition of these plates allows for the differential detection of uropathogenic microorganisms. For plating on ciprofloxacin, plates containing ciprofloxacin at 0.5, 2 or 10 mg/L were prepared using HiChromeTM UTI Agar (HiMedia Laboratories Pvt, Ltd., India). Sample was plated using standard quadrant plating technique[2]. The rest of the sample was split into two tubes (with and without 10% glycerol) and stored at -80°C. The plates were incubated at 37°C for 16-20 hours, inspected visually, and the growth characteristics were documented including the quantitative growth level of potential *E. coli*, Gram-negative and total bacteria. Single colonies with *E. coli*-like morphology were isolated from all growth-positive CIP plates and a random selection of UTI plates for further analysis. The mixed cultures were then preserved in 10% glycerol-containing freezer medium and stored at -80°C.

#### Quantification of relative *E. coli* and CIP-resistant *E. coli* abundance in fecal samples

As described above, the semi-quantitative quadrant streaking was performed on chromogenic UTI agar to estimate the relative abundance of *E. coli* (on plain agar plates) and CIP-resistant *E. coli* (on ciprofloxacin-supplemented plates) in fecal samples. Colony counts were taken from the final growth quadrant, where individual colonies could be clearly distinguished. Morphological characteristics provided by the manufacturer were used to differentiate *E. coli* from other Gram-negative and Gram-positive bacteria (see **Supplemental Figure S3A** for example). This approach allowed calculation of the proportion of *E. coli* colonies relative to the total Gram-negative or overall bacterial growth. The abundance of CIP-resistant *E. coli* relative to total *E. coli* was determined by comparing colony counts on ciprofloxacin-containing versus plain agar.

#### Identification of TMP/STX-R and 3GC-R *E. coli* in fecal samples

An aliquot of every fecal sample stored in glycerol was resuspended in 100 µL of Mueller-Hinton (MH) broth, incubated at 37°C for 2 hours, plated on UTI agar supplemented with ciprofloxacin (CIP, 0.5 mg/L), trimethoprim/sulfamethoxazole (TMP/STX, 4/76 mg/L), ceftazidime (CAZ, 8 mg/L), cefotaxime (CTX, 2 mg/L), and without antibiotics. After overnight incubation plates were processed same way as described above. Growth on UTI-CIP plate served as data reproducibility control. All potentially resistant *E. coli* colonies were subcultured on MH-agar supplemented with respective antibiotic to confirm their non-susceptibility status.

#### Clinical urine sample collection and processing

If a study participant submitted a urine sample to KPWA clinical laboratory, a routine urinalysis test was performed, followed by culture and sensitivity testing if required. Starting February 2022, the UW laboratory was provided an aliquot of the urine sample, which was processed using the same protocol as fecal sample described above, with initial plating on plain and CIP-UTI plates. Potential *E. coli* were saved as at least 4-5 individual colonies and used for antibiotic susceptibility testing and identification of *E. coli* clonality (see below). The time lapse between the beginning of the study (May 2021) and the start of availability of clinical urine samples to the UW lab accounts for the lack of clonal information for some *E. coli*-caused UTIs.

#### Identification of *E. coli* clonality

*E. coli* clonality was determined by CH typing based on *fumC*/*fimH* sequencing[3], presence of QRDR mutations was determined by sequencing of *gyrA* and *parC*[4]. All reactions were carried out by 2-step colony PCR. Briefly, a single *E. coli* colony was resuspended in 50 µL of sterile water and heated at 98°C for 10 min. Primary PCR reactions were set up in a 15 µL volume using DreamTaq Mastermix (Thermofisher, USA), supplemented with 0.5 µM forward and reverse primers, and 1.5 µL of the boiled colony template. Primary PCR was run for 30 cycles under the manufacturer's recommended conditions. Subsequently, 1 µL of the PCR1 product was used for an additional PCR reaction, using nested forward and reverse primers supplemented with T7 and T7-Term tails, respectively. The nested PCR was run for 15 cycles under the same conditions, aiming to obtain a highly specific single band with T7-tailed primers suitable for downstream sequencing. The primer sequences can be found in **Supplemental Table S4**.

#### Testing antibiotic resistance of *E. coli* isolates

Resistance of fecal *E. coli* isolates to antibiotics of interest was performed using agar dilution method as described in CLSI manual [5]. Resistance of urinary *E. coli* isolates to a panel of 12 antibiotics was tested using Kirby-Bauer disk diffusion method as described in CLSI manual [5].

#### Metagenomic analysis

*Sequencing*.

A subset of 36 pairs of fecal and clinical urine samples was analyzed to assess the relative abundance of *E. coli* among enterobacteria, as well as the presence and prevalence of specific *E. coli* clones. Different dilutions of samples were plated on MacConkey agar to obtain $10^3$-$10^4$ colonies per plate. Total growth was pooled and DNA was extracted from pooled colonies. DNA was used for shotgun metagenomic sequencing on an Illumina MiSeq platform using the MiSeq 600 cycle v3 kit, following the manufacturer's guidelines. Genomic DNA libraries were prepared with the Nextera XT Library Prep Kit (Illumina, CA). The raw reads were analyzed for species composition and urinary *E. coli* clone abundance as follows.

*Determining species composition in sample*.

Shotgun metagenomic reads were analyzed using the PATRIC Taxonomic Profiling Tool (https://patricbrc.org/). Raw reads were quality-checked with FastQC and trimmed using Trim Galore (Phred < 20). High-quality reads were processed through PATRIC, which employs Kraken2 for taxonomic classification and Bracken for abundance estimation. Taxonomic profiles, including relative species abundances, were generated and exported for downstream analysis.

*Detection and quantification of urinary E. coli clone in samples*.

The presence and abundance of the urinary *E. coli* clone in fecal samples were determined through comparative genomics and targeted read alignment. First, the sequenced urinary clone was compared to genomes of the same sequence type (ST) and different STs using both in-house sequenced isolates and publicly available genomes from EnteroBase (https://enterobase.warwick.ac.uk/). This analysis identified ST-specific alleles and isolate-specific SNPs unique to the clone of interest.

Raw sequencing reads were aligned to the identified alleles using BWA-MEM with default parameters. The resulting SAM file was converted to a sorted and indexed BAM file using SAMtools for efficient variant calling and read depth analysis. Variant calling was performed using BCFtools mpileup, with a maximum depth (-d 250), a minimum mapping quality (-q 60), and a minimum base quality (-Q 30), ensuring high-confidence variant detection. The bcftools call function was used in multiallelic (-m) and variant-only

(-v) mode, with a ploidy setting of 1 (haploid), to generate a compressed VCF file containing identified variants. The VCF file was then indexed and queried to extract depth of coverage (%DP) and allele frequencies (%AD) for each position of interest.

The output was manually examined for the presence of alleles unique to the strain of interest. The relative abundance of the strain was calculated as the proportion of sequencing reads supporting the unique SNPs, using the following formula:

*Abundance (%) = ∑ RefReads (unique SNPs) / ∑ (RefReads (unique SNPs) + AltReads (unique SNPs)) × 100*

If no unique SNPs were detected, the alignment process was repeated for other identified alleles to confirm the absence of the strain in the sample.

*Assessment of E. coli clonal diversity in fecal samples by metagenomics*

The number of distinct *E. coli* CH types (defined by *fumC/fimH* allele combination) present in fecal samples was estimated via two complementary approaches. In the first, metagenomic reads from 14 fecal samples were aligned to a curated reference database of *fumC* and *fimH* alleles using BWA-MEM with the same parameters described above for urinary *E. coli* clone detection. Variant calling was performed using BCFtools mpileup, and allele frequencies at polymorphic positions within *fumC* and *fimH* were used to model the number of distinct CH types present in each sample using the same abundance formula described above. This analysis was limited by the sequencing depth achieved (~100× average read depth per base), which may constrain reliable detection of minor clonal subpopulations.

In the second approach, 29 fecal samples carrying the ST131-*H*30 clone were selected to compare clonal diversity with UTI occurrence during follow-up. Samples were matched by age and fecal *E. coli* abundance and included equal proportions of enrollees who did and did not develop a UTI during follow-up. DNA purified from frozen stocks of bacterial growth recovered from fecal samples on UTI-selective plates was analyzed using the Population-Level Allele Profiler (PLAP) method developed in our laboratory [6]. Briefly, *fumC* and *fimH* genes were amplified by PCR and amplicons were sequenced using Plasmidsaurus (Oxford Nanopore Technology;

https://plasmidsaurus.com) and analyzed to determine the number of distinct CH clonotypes per sample. As with the metagenomic approach, this analysis was limited by the use of selective plate growth rather than total fecal DNA, which may underrepresent minor *E. coli* subpopulations present at low abundance. For five ST131-H30 fecal carrying samples both amplicon and metagenomics sequencing identified same number of clones, confirming the robustness of this method.

#### Comparison of paired fecal and clinical urine *E. coli*

*Whole Genome Sequencing*.

Whole genome sequencing was performed on an Illumina MiSeq platform using the MiSeq 600 cycle v3 kit, following the manufacturer's guidelines. Genomic DNA libraries were prepared with the Nextera XT Library Prep Kit (Illumina, CA). Raw data were uploaded to the Enterobase database (https://enterobase.warwick.ac.uk/) for genome assembly, allele calling, and wgMLST and cgMLST assignments (Enterobase uberstrain names are listed in Supplemental Data).

*Selection of reference genomes for phylogenetic analysis*

For each fecal-urinary isolate pair, the uropathogenic isolate was assigned an Achtman MLST sequence type (ST), *fimH*, *gyrA*, and *parC* allele numbers via EnteroBase. The EnteroBase database was then searched for publicly available genomes matching the same ST-*fimH*-*gyrA*-*parC* combination. cgMLST allelic profiles were downloaded for all matching strains; for over-represented STs, the dataset was filtered by isolation year (after 2010), source (human), and/or geographic origin (North America) to reduce redundancy while retaining representativeness. The number of matching genomes ranged from 18 (ST450, isolate ID15727) to 3,000 (ST1193), with 100–200 genomes retrieved on average. The corresponding fecal isolate was always included in the dataset.

To identify the genomes most closely related to each uropathogenic isolate, a custom R script (**Supplemental R script**) was applied to the combined cgMLST allelic profiles. Briefly, allelic profiles of the uropathogen and all reference genomes were consolidated and restricted to loci present in the focal pair. A SNP-based alignment was constructed by resolving each allele call to its nucleotide sequence at variable cgMLST loci using a curated allele FASTA database, retaining only positions that were polymorphic across the dataset and for which the query allele sequence was available. Pairwise SNP distances between each reference genome and the uropathogen (query) were computed from this alignment. Genomes with identical SNP profiles were deduplicated, retaining one representative. Up to 15 reference genomes with the smallest SNP distance to the query were selected; however, for rare STs with limited public representation, fewer genomes were available — for example, only 3 genomes were identified for ST1905 (ID13476) and ST450 (ID15727), and 5 for ST5308 (ID11255). The selected reference genomes, together with the matched fecal isolate, constituted the final dataset for phylogenetic tree construction. The resulting SNP alignment was used as input for maximum likelihood phylogenetic inference in MEGA12.

*Plasmid analysis of discordant fecal-urinary E. coli pairs*

To assess whether plasmid dynamics could explain resistance discordance between fecal and urinary *E. coli* isolates, plasmid content was analyzed for a subset of 20 discordant pairs in which both isolates were available for whole-genome sequencing. Resistance genes were identified using ***ResFinder*** and plasmid replicon types were determined using ***PlasmidFinder***, both available through the ***Center for Genomic Epidemiology (CGE)*** platform (https://genomicepidemiology.org), using default parameters and the Enterobacteriaceae database. Contig IDs carrying resistance genes in ***ResFinder*** were cross-referenced with plasmid-positive contigs identified by ***PlasmidFinder*** to determine whether resistance genes were plasmid-borne.

#### Supplemental Tables

**Table S1.** Clonotype distribution of multidrug-resistant CIP-R *E. coli* isolated from fecal samples. ST, sequence type, CC, clonal complex, H, *fimH* allele, Q, number of QRDR mutations in *gyrA* and/or *parC* known to confer CIP-resistance; for ST69-H27(2Q) and ST38-H5(1Q) and ST38-H65(3Q) *gyrA* and *parC* allele numbers are listed after comma to indicate different CIP-R clones.

| CIP-R E. coli Clone | No. CIPR-SC (%Total) | No. CIPR MDR SC (%Total) |
|---|---|---|
| H30 | 70 (20.3) | 27 (18.4) |
| ST1193 | 68 (19.7) | 29 (19.7) |
| ST131-H41(1Q) | 15 (4.3) | 8 (5.4) |
| ST69-H27, 14-76(2Q) | 15 (4.3) | 4 (2.7) |
| ST69-H27, 14-13(2Q) | 12 (3.5) | 6 (4.1) |
| ST648-H0(3Q) | 9 (2.6) | 5 (3.4) |
| ST69-H27(0Q) | 8 (2.3) | 3 (2) |
| ST69-H27(1Q) | 6 (1.7) | 3 (2) |
| ST69-H27(3Q) | 5 (1.4) | 3 (2) |
| ST38-H5(3Q) | 4 (1.2) | 3 (2) |
| ST10-H27(1Q) | 4 (1.2) | 2 (1.4) |
| ST354-H58(4Q) | 3 (0.9) | 3 (2) |
| ST636-H0(1Q) | 3 (0.9) | 3 (2) |
| ST394-H30(1Q) | 3 (0.9) | 2 (1.4) |
| ST95-H41(1Q) | 3 (0.9) | 1 (0.7) |
| ST38-H65, 11-14(3Q) | 3 (0.9) | 0 (0) |
| ST405-H27(3Q) | 2 (0.6) | 2 (1.4) |
| ST44(CC10)-H54(3Q) | 2 (0.6) | 2 (1.4) |
| ST6151(CC21)-H0(3Q) | 2 (0.6) | 2 (1.4) |
| ST773(CC10)-H0(3Q) | 2 (0.6) | 2 (1.4) |
| ST10-H54(4Q) | 2 (0.6) | 1 (0.7) |
| ST38-H65, 11-11(3Q) | 2 (0.6) | 1 (0.7) |
| ST457-H145(3Q) | 2 (0.6) | 1 (0.7) |
| ST648-H30(3Q) | 2 (0.6) | 1 (0.7) |
| ST6754(CC101)-H0(1Q) | 2 (0.6) | 1 (0.7) |
| ST69(3Q) | 2 (0.6) | 1 (0.7) |
| CC10-H27(3Q) | 2 (0.6) | 0 (0) |
| ST10-H215(3Q) | 2 (0.6) | 0 (0) |
| ST1177(CC38)-H65(2Q) | 2 (0.6) | 0 (0) |
| ST224(CC58)-H61(3Q) | 2 (0.6) | 0 (0) |

| | | |
|---|---|---|
| ST38-H5, 14-36(1Q) | 2 (0.6) | 0 (0) |
| ST48(CC10)-H34(0Q) | 2 (0.6) | 0 (0) |
| ST93(CC10)-H30(3Q) | 2 (0.6) | 0 (0) |
| CC10-H54(3Q) | 1 (0.3) | 1 (0.7) |
| CC349-H54(1Q) | 1 (0.3) | 1 (0.7) |
| CH12-342(0Q) | 1 (0.3) | 1 (0.7) |
| ST10-H27(0Q) | 1 (0.3) | 1 (0.7) |
| ST10-H28(1Q) | 1 (0.3) | 1 (0.7) |
| ST117-H97(3Q) | 1 (0.3) | 1 (0.7) |
| ST127-H2(1Q) | 1 (0.3) | 1 (0.7) |
| ST12-H7(0Q) | 1 (0.3) | 1 (0.7) |
| ST131-H41(4Q) | 1 (0.3) | 1 (0.7) |
| ST1380(CC394)-H1057(0Q) | 1 (0.3) | 1 (0.7) |
| ST14-H27(1Q) | 1 (0.3) | 1 (0.7) |
| ST1844(CC111)-H38(0Q) | 1 (0.3) | 1 (0.7) |
| ST2006(CC58)-H61(3Q) | 1 (0.3) | 1 (0.7) |
| ST2197(CC10)-H23(3Q) | 1 (0.3) | 1 (0.7) |
| ST224(CC58)-H54(3Q) | 1 (0.3) | 1 (0.7) |
| ST362-H96(0Q) | 1 (0.3) | 1 (0.7) |
| ST38-H5, 294-36(1Q) | 1 (0.3) | 1 (0.7) |
| ST38-H54(1Q) | 1 (0.3) | 1 (0.7) |
| ST38-H65, 11-13(3Q) | 1 (0.3) | 1 (0.7) |
| ST393(CC69)-H54(3Q) | 1 (0.3) | 1 (0.7) |
| ST457-H145(0Q) | 1 (0.3) | 1 (0.7) |
| ST58-H1325(1Q) | 1 (0.3) | 1 (0.7) |
| ST617(CC10)-H29(3Q) | 1 (0.3) | 1 (0.7) |
| ST62-H44(0Q) | 1 (0.3) | 1 (0.7) |
| ST636-H27(1Q) | 1 (0.3) | 1 (0.7) |
| ST69-H25(1Q) | 1 (0.3) | 1 (0.7) |
| ST69-H27, 14-2(2Q) | 1 (0.3) | 1 (0.7) |
| ST73-H10(1Q) | 1 (0.3) | 1 (0.7) |
| ST95-H41(0Q) | 1 (0.3) | 1 (0.7) |
| ST95-H468(1Q) | 1 (0.3) | 1 (0.7) |
| ST95-H99(1Q) | 1 (0.3) | 1 (0.7) |
| CC10-H34(3Q) | 1 (0.3) | 0 (0) |
| CC2797-H1637(0Q) | 1 (0.3) | 0 (0) |
| CC349-H54(3Q) | 1 (0.3) | 0 (0) |
| CC58-H121(1Q) | 1 (0.3) | 0 (0) |
| CH12-1580(1Q) | 1 (0.3) | 0 (0) |
| CH14-27(0Q) | 1 (0.3) | 0 (0) |
| ST10-H0(0Q) | 1 (0.3) | 0 (0) |
| ST10-H28(0Q) | 1 (0.3) | 0 (0) |
| ST10-H30 | 1 (0.3) | 0 (0) |

| | | |
|---|---|---|
| ST10-H54(0Q) | 1 (0.3) | 0 (0) |
| ST10-H54(3Q) | 1 (0.3) | 0 (0) |
| ST1163 | 1 (0.3) | 0 (0) |
| ST131-H41(3Q) | 1 (0.3) | 0 (0) |
| ST140(CC95)-H15(1Q) | 1 (0.3) | 0 (0) |
| ST1431(CC58)-H32(3Q) | 1 (0.3) | 0 (0) |
| ST155(CC58)-H0(0Q) | 1 (0.3) | 0 (0) |
| ST1723-H38(3Q) | 1 (0.3) | 0 (0) |
| ST206(CC77)-H0(0Q) | 1 (0.3) | 0 (0) |
| ST206(CC77)-H23(3Q) | 1 (0.3) | 0 (0) |
| ST216-H69(1Q) | 1 (0.3) | 0 (0) |
| ST2772(CC58)-H26(0Q) | 1 (0.3) | 0 (0) |
| ST2973-H31(2Q) | 1 (0.3) | 0 (0) |
| ST349-H54(0Q) | 1 (0.3) | 0 (0) |
| ST372-H1639(1Q) | 1 (0.3) | 0 (0) |
| ST3863(CC21)-H1632(0Q) | 1 (0.3) | 0 (0) |
| ST38-H0(3Q) | 1 (0.3) | 0 (0) |
| ST38-H30(3Q) | 1 (0.3) | 0 (0) |
| ST38-H5(0Q) | 1 (0.3) | 0 (0) |
| ST405-H29(3Q) | 1 (0.3) | 0 (0) |
| ST4267(CC58)-H54(0Q) | 1 (0.3) | 0 (0) |
| ST4305(CC10)-H0(2Q) | 1 (0.3) | 0 (0) |
| ST450-H34(3Q) | 1 (0.3) | 0 (0) |
| ST452-H0(0Q) | 1 (0.3) | 0 (0) |
| ST5150(CC38)-H65(2Q) | 1 (0.3) | 0 (0) |
| ST569-H5(1Q) | 1 (0.3) | 0 (0) |
| ST5869(CC3945)-H31(3Q) | 1 (0.3) | 0 (0) |
| ST58-H30(0Q) | 1 (0.3) | 0 (0) |
| ST58-H32(0Q) | 1 (0.3) | 0 (0) |
| ST59-H34(0Q) | 1 (0.3) | 0 (0) |
| ST68-H382(1Q) | 1 (0.3) | 0 (0) |
| ST69(0Q) | 1 (0.3) | 0 (0) |
| ST744(CC10)-H54(3Q) | 1 (0.3) | 0 (0) |
| ST744-H54(3Q) | 1 (0.3) | 0 (0) |
| ST80-H1(0Q) | 1 (0.3) | 0 (0) |
| ST8492(CC58)-H38(3Q) | 1 (0.3) | 0 (0) |
| ST906(CC58)-H32(0Q) | 1 (0.3) | 0 (0) |
| ST93(CC10)-H0(0Q) | 1 (0.3) | 0 (0) |
| ST969(CC625)-H115(1Q) | 1 (0.3) | 0 (0) |
| not determined | 1 (0.3) | 0 (0) |
| Total CIP-R SC | 345 | 147 |

**Table S2**. UTI *E. coli* resistant to antibiotics.

| ID | ET [a] | UTI *E. coli* clone [b] | Resistance [c] | CU=F [d] |
|---|---|---|---|---|
| 10094 | 7 | sample unavailable | TS | YES |
| 10152 | 17 | ST131-H30 | CIP | **no** |
| 10226 | 13 | ST1193 | CIP TS | YES |
| 10273 | 2 | sample unavailable | TS | YES |
| 10571 | 5 | sample unavailable | TS 3GC | YES |
| 10860 | 6 | sample unavailable | TS | **no** |
| 10891 | 2 | sample unavailable | CIP | YES |
| 11186 | 14 | ST69-H27 | TS | YES |
| 11212 | 14 | ST963(CC38)-H26 | 3GC | YES |
| 11255 | 9 | ST5308(CC674)-H233 | TS | YES |
| 11403 | 4 | sample unavailable | 3GC | **no** |
| 11595 | 17 | ST73-H10 | TS | YES |
| 11921 | 7 | sample unavailable | TS | YES |
| 12118 | 18 | ST404(CC14)-H27 | TS | YES |
| 12211 | 15 | ST131-H30 (0 QRDR) | TS | YES |
| 12247 | 2 | ST131-H30 | CIP TS 3GC | YES |
| 12450 | 1 | sample unavailable | CIP | YES |
| 12736 | 9 | ST131-H30 | CIP | YES |
| 12815 | 18 | ST404(CC14)-H27 | TS | YES |
| 13026 | 7 | ST131-H30 | TS | YES |
| 13168 | 11 | ST69-H27(2Q) | CIP TS | YES |
| 13561 | 16 | ST131-H30 | CIP | **no** |
| 13792 | 9 | ST131-H30 | CIP | YES |
| 13844 | 6 | ST963(CC38)-H26 | 3GC | YES |
| 14315 | 1 | sample unavailable | CIP | YES |
| 14440 | 4 | sample unavailable | CIP | YES |
| 14509 | 11 | ST131-H30 | CIP | YES |
| 14737 | 3 | sample unavailable | CIP | YES |
| 15034 | 17 | ST131-H41 | TS | YES |
| 15727 | 6 | CC10-H54 | CIP TS | YES |
| 15823 | 13 | ST69-H27 | TS | **no** |
| 15924 | 7 | ST131-H30 | CIP | YES |
| 16419 | 17 | ST1193 | CIP TS | YES |
| 16572 | 9 | ST69-H27(2Q) | CIP | YES |
| 16660 | 12 | ST131-H30 | CIP | YES |
| 16663 | 17 | ST131-H41 | CIP | YES |

[a] ET, time elapsed between UTI and fecal sample, in months.

[b] No sample – sample not available in UW laboratory, clonal typing not performed.

[c] Resistance of uropathogenic *E. coli* was determined in KPWA clinical laboratory and confirmed in UW laboratory if sample was available. CIP, ciprofloxacin, TS, trimethoprim/sulfamethoxazole, 3GC, 3rd generation cephalosporins (ceftazidime and/or ceftriaxone)

[d] CU=F, clinical uropathogenic *E. coli* clone was identified among fecal *E. coli* from the same participant

**Table S3**. Non-*E. coli* UTI uropathogens.

| ID | ET [a] | Urinary pathogen [b] | Resistance [b] | Fecal *E. coli* Resistance |
|---|---|---|---|---|
| 11653 | 3 | *C. koseri* (sample unavailable) | none | CIP |
| 12140 | 3 | *K. pneumoniae* (sample unavailable) | none | none |
| 10700 | 4 | *K. pneumoniae* (sample unavailable) | none | none |
| 12218 | 7 | *E. faecalis* | none | none |
| 11544 | 8 | *K. pneumoniae* (sample unavailable) | none | none |
| 15712 | 11 | *C. freundii* | none | TMP/SXT |
| 10430 | 11 | *Klebsiella sp.* | none | none |
| 13734 | 11 | *K. pneumoniae* | none | none |
| 15321 | 12 | *P. mirabilis* | none | none |
| 14115 | 14 | *K. pneumoniae* | none | none |
| 11764 | 14 | *P. mirabilis* | none | none |
| 12805 | 15 | *K. pneumoniae* | none | none |
| 13446 | 15 | *P. mirabilis* | none | none |
| 15189 | 16 | *Klebsiella sp.* | none | TMP/SXT |
| 12041 | 17 | *K. pneumoniae* (sample unavailable) | none | none |
| 10944 | 17 | *Salmonella sp.* | none | none |

[a] ET, time elapsed between UTI and fecal sample, in months.

[b] Both uropathogen's species and resistance were determined in KPWA clinical laboratory and confirmed in UW laboratory if sample was available.

**Supplemental Table S4**. Primers used for CH typing, *gyrA-parC* sequencing.

| **Test** | **Target** | **Primer name** | **Sequence** | **Ref** |
|---|---|---|---|---|
| **CH typing** | **fumC: PCR1** | fumC-F | GCATCACAGGTCGCCAGCG | This study |
| | | fumC-R | GTACGCAGCGAAAAAGATTC | |
| | **fumC: Nested** | fumC-F'-T7Pro | TAATACGACTCACTATAGGGGCGCTTCAAATTTGTTCGG | This study |
| | | fumC-R'-T7Term | GCTAGTTATTGCTCAGCGGGTACGCAGCGAAAAAGATTC | |
| | **fimH: PCR1** | fimH-F | CTGTTTGCTGTACTGCTGATG | This study |
| | | fimH-R | CCACAATAAACGGTAAGAGGAAT | |
| | **fimH: Nested** | fimH-F'-T7Pro | TAATACGACTCACTATAGGGACTGCTGATGGGCTGGTC | This study |
| | | fimH-R'-T7Term | GCTAGTTATTGCTCAGCGGAGGAATTGGCACTGAACC | |
| **Detection of QRDR SNPs** | **gyrA: PCR1** | gyrA-F | CGACCTTGCGAGAGAAAT | This study |
| | | gyrA-R | GTTCCATCAGCCCTTCAA | |
| | **gyrA: Nested** | gyrA-F'-T7Pro | TAATACGACTCACTATAGGGCGAGAGAAATTACACCG | This study |
| | | gyrA-R'-T7Term | GCTAGTTATTGCTCAGCGGAGCCCTTCAATGCT | |
| | **parC: PCR1** | parC-F | CGATTGCCGCCTGAGCCACTT | This study |
| | | parC-R | GCGAATAAGTTGAGGAATCAG | |
| | **parC: Nested** | parC-F'-T7Pro | TAATACGACTCACTATAGGGTGAGCCACTTCACGCA | This study |
| | | parC-R'-T7Term | GCTAGTTATTGCTCAGCGGGAGGAATCAGAATTAA | |

**Table S5. Classification of participants with diagnosed UTI by uropathogen and baseline fecal *E. coli* carriage**

| Days between Fecal sample and UTI | Uropathogen [a] | Uropathogen's Resistance to: | | | UTI sample available for clonal typing | Clonally matched Uropathogen *E. coli* found in fecal sample | ST131-*H*30 in fecal sample | ST1193 in fecal sample | Study ID |
|---|---|---|---|---|---|---|---|---|---|
| | | CIP | TMP/SXT | 3GC | | | | | |
| 3 | *E. coli* | - | - | - | no | . | no | no | 10632 |
| 3 | *E. coli* | - | - | - | no | . | no | no | 13754 |
| 4 | *E. coli* | + | - | - | no | . | YES | no | 14315 |
| 6 | *E. coli* | + | - | - | no | . | YES | no | 12450 |
| 9 | *E. coli* | - | - | - | no | . | no | no | 10973 |
| 10 | *E. coli* | - | - | - | no | . | no | no | 13471 |
| 13 | . | . | . | . | . | . | no | no | 12366 |
| 15 | *E. coli* | - | - | - | no | . | no | no | 13555 |
| 20 | *E. coli* | - | - | - | no | . | no | no | 12254 |
| 20 | *E. coli* | - | - | - | no | . | no | no | 14779 |
| 21 | *E. coli* | - | - | - | no | . | no | no | 10345 |
| 30 | *E. coli* | - | - | - | no | . | no | no | 12384 |
| 32 | *E. coli* | - | - | - | no | . | no | no | 12684 |
| 38 | *E. coli* | - | + | - | no | . | no | no | 10273 |
| 40 | . | . | . | . | . | . | no | no | 13707 |
| 48 | *E. coli* | - | - | - | no | . | no | no | 15906 |
| 49 | *E. coli* | + | + | + | YES | YES | YES | no | 12247 |
| 55 | *E. coli* | - | - | - | no | . | no | no | 13958 |
| 59 | . | . | . | . | . | . | no | YES | 14492 |
| 61 | *E. coli* | + | - | - | no | . | no | YES | 10891 |
| 68 | . | . | . | . | . | . | no | no | 15761 |
| 68 | . | . | . | . | . | . | no | no | 16653 |
| 70 | . | . | . | . | . | . | no | no | 16348 |
| 75 | *C. koseri* | - | - | - | no | . | no | no | 11653 |
| 78 | *E. coli* | - | - | - | no | . | no | no | 12763 |
| 80 | *E. coli* | - | - | - | no | . | no | no | 12813 |
| 81 | *E. coli* | - | - | - | no | . | no | no | 14732 |
| 85 | . | . | . | . | . | . | no | no | 15101 |
| 86 | *E. coli* | - | - | - | no | . | no | no | 14145 |
| 86 | *E. coli* | + | - | - | no | . | no | YES | 14737 |
| 86 | . | . | . | . | . | . | no | YES | 14944 |
| 86 | . | . | . | . | . | . | no | no | 16471 |
| 93 | *K. pneumoniae* | - | - | - | no | . | no | no | 12140 |
| 98 | . | . | . | . | . | . | no | no | 13473 |
| 102 | . | . | . | . | . | . | no | no | 15011 |
| 104 | . | . | . | . | . | . | no | no | 14822 |
| 107 | *E. coli* | + | - | - | no | . | no | no | 14440 |
| 108 | . | . | . | . | . | . | no | no | 11476 |
| 112 | *E. coli* | - | - | - | no | . | no | no | 10157 |
| 115 | *K. pneumoniae* | - | - | - | no | . | no | no | 10700 |
| 116 | *E. coli* | - | - | + | no | . | no | no | 11403 |
| 117 | *E. coli* | - | - | - | no | . | no | no | 11520 |
| 119 | *E. coli* | - | - | - | no | . | no | no | 14406 |
| 125 | *E. coli* | - | - | - | no | . | no | no | 10473 |
| 128 | *E. coli* | - | - | - | no | . | no | no | 12025 |
| 129 | . | . | . | . | . | . | no | no | 12720 |
| 131 | *E. coli* | - | - | - | no | . | no | no | 10644 |

Table S5. Classification of participants with diagnosed UTI by uropathogen and baseline fecal *E. coli* carriage

| Days between Fecal sample and UTI | Uropathogen [a] | Uropathogen's Resistance to: | | | UTI sample available for clonal typing | Clonally matched Uropathogen *E. coli* found in fecal sample | ST131-*H*30 in fecal sample | ST1193 in fecal sample | Study ID |
|---|---|---|---|---|---|---|---|---|---|
| | | CIP | TMP/SXT | 3GC | | | | | |
| 131 | *E. coli* | - | - | + | YES | YES | YES | no | 11212 |
| 140 | . | . | . | . | . | . | no | no | 12420 |
| 146 | *E. coli* | - | - | - | no | . | no | no | 12354 |
| 146 | *E. coli* | - | - | - | YES | no | no | no | 16385 |
| 147 | *E. coli* | - | + | + | no | . | no | no | 10571 |
| 148 | . | . | . | . | . | . | no | no | 10740 |
| 178 | . | . | . | . | . | . | no | no | 14606 |
| 181 | *E. coli* | - | - | + | YES | YES | no | no | 13844 |
| 183 | *E. coli* | - | - | - | YES | YES | no | no | 16608 |
| 186 | *E. coli* | - | + | - | no | . | no | no | 10860 |
| 186 | . | . | . | . | . | . | no | no | 14043 |
| 187 | *E. coli* | - | - | - | no | . | no | no | 11582 |
| 188 | *E. coli* | - | + | - | no | . | no | no | 11921 |
| 192 | *E. coli* | - | + | - | no | . | no | no | 10094 |
| 193 | *E. coli* | + | - | - | YES | YES | no | no | 15727 |
| 194 | . | . | . | . | . | . | YES | no | 13037 |
| 198 | . | . | . | . | . | . | no | no | 11841 |
| 199 | *E. coli* | - | - | - | YES | no | no | no | 12262 |
| 202 | *E. coli* | - | - | - | YES | no | no | no | 13062 |
| 204 | *E. coli* | + | - | - | YES | YES | YES | no | 15924 |
| 207 | . | . | . | . | . | . | no | no | 12248 |
| 208 | *E. coli* | - | + | - | YES | YES | no | no | 13026 |
| 209 | *E. coli* | - | - | - | no | . | no | no | 15862 |
| 210 | *E. faecalis* | - | - | - | no | . | no | no | 12218 |
| 214 | *E. coli* | - | - | - | YES | YES | no | no | 14053 |
| 221 | *E. coli* | - | - | - | YES | no | no | no | 15616 |
| 224 | *E. coli* | - | - | - | YES | YES | no | no | 14907 |
| 225 | *K. pneumoniae* | - | - | - | no | . | no | no | 11544 |
| 227 | *E. coli* | - | - | - | YES | YES | no | no | 13079 |
| 227 | *E. coli* | - | - | - | YES | YES | no | no | 14588 |
| 233 | . | . | . | . | . | . | no | no | 11031 |
| 239 | *E. coli* | - | - | - | no | . | no | no | 10480 |
| 241 | *E. coli* | - | - | - | YES | no | no | no | 12858 |
| 243 | . | . | . | . | . | . | no | no | 13122 |
| 249 | . | . | . | . | . | . | no | no | 14604 |
| 253 | *E. coli* | - | + | - | YES | YES | no | no | 11255 |
| 257 | *E. coli* | - | - | - | YES | no | no | no | 10641 |
| 259 | *E. coli* | - | - | - | YES | no | no | no | 15249 |
| 264 | *E. coli* | + | - | - | YES | YES | no | no | 16572 |
| 269 | *E. coli* | - | - | - | YES | YES | no | no | 13476 |
| 269 | *E. coli* | - | - | - | YES | no | no | no | 13975 |
| 270 | . | . | . | . | . | . | no | no | 15904 |
| 271 | *E. coli* | + | - | - | YES | YES | YES | no | 13792 |
| 272 | . | . | . | . | . | . | no | no | 12427 |
| 276 | . | . | . | . | . | . | no | no | 13684 |
| 279 | *E. coli* | - | - | - | YES | YES | YES | no | 11892 |
| 282 | *E. coli* | + | + | + | YES | YES | YES | no | 12736 |

Table S5. Classification of participants with diagnosed UTI by uropathogen and baseline fecal *E. coli* carriage

| Days between Fecal sample and UTI | Uropathogen [a] | Uropathogen's Resistance to: | | | UTI sample available for clonal typing | Clonally matched Uropathogen *E. coli* found in fecal sample | ST131-*H*30 in fecal sample | ST1193 in fecal sample | Study ID |
|---|---|---|---|---|---|---|---|---|---|
| | | CIP | TMP/SXT | 3GC | | | | | |
| 284 | *E. coli* | - | - | - | YES | YES | no | no | 11235 |
| 288 | *E. coli* | - | - | - | YES | no | no | no | 15035 |
| 292 | *E. coli* | + | + | - | YES | YES | no | YES | 10226 |
| 293 | . | . | . | . | . | . | no | no | 12986 |
| 302 | *E. coli* | - | - | - | YES | no | no | no | 14534 |
| 303 | *E. coli* | - | - | - | YES | no | no | no | 12988 |
| 322 | . | . | . | . | . | . | no | no | 15399 |
| 325 | *E. coli* | - | - | - | YES | no | no | no | 12635 |
| 330 | *E. coli* | + | - | - | YES | YES | YES | no | 14509 |
| 331 | . | . | . | . | . | . | no | no | 13906 |
| 332 | *Citrobacter sp.* | - | - | - | no | . | no | no | 15712 |
| 333 | . | . | . | . | . | . | no | no | 15401 |
| 338 | *E. coli* | + | + | - | YES | YES | no | no | 13168 |
| 338 | . | . | . | . | . | . | no | no | 13300 |
| 338 | *K. pneumoniae* | - | - | - | no | . | no | no | 13734 |
| 339 | . | . | . | . | . | . | no | no | 12067 |
| 340 | *E. coli* | - | - | - | YES | YES | no | no | 11818 |
| 341 | *E. coli* | - | - | - | YES | no | no | no | 13371 |
| 345 | . | . | . | . | . | . | no | no | 14924 |
| 347 | *Klebsiella sp.* | - | - | - | no | . | no | no | 10430 |
| 347 | *E. coli* | - | - | - | YES | YES | no | no | 12316 |
| 347 | *E. coli* | + | - | - | YES | YES | YES | no | 16660 |
| 348 | *E. coli* | - | - | - | YES | no | no | no | 13018 |
| 348 | . | . | . | . | . | . | no | no | 16402 |
| 350 | *P. mirabilis* | - | - | - | no | . | no | no | 15321 |
| 359 | *E. coli* | - | - | - | YES | YES | YES | no | 13363 |
| 367 | *E. coli* | - | - | - | YES | no | no | no | 12959 |
| 369 | *E. coli* | - | + | - | YES | no | no | no | 15823 |
| 376 | *E. coli* | - | - | - | YES | YES | no | no | 12547 |
| 377 | *E. coli* | - | - | - | no | . | no | no | 12048 |
| 378 | *E. coli* | - | - | - | YES | YES | no | no | 10148 |
| 382 | *E. coli* | - | - | - | YES | no | no | no | 10048 |
| 391 | *E. coli* | - | - | - | YES | no | YES | no | 14510 |
| 395 | *E. coli* | - | - | - | no | . | no | no | 13323 |
| 395 | . | . | . | . | . | . | no | no | 16598 |
| 399 | *E. coli* | - | - | - | YES | no | YES | no | 16344 |
| 413 | *P. mirabilis* | - | - | - | no | . | no | no | 11764 |
| 416 | *K. pneumoniae* | - | - | - | no | . | no | no | 14115 |
| 423 | . | . | . | . | . | . | no | YES | 14239 |
| 425 | *E. coli* | - | - | - | no | . | no | no | 15480 |
| 431 | *E. coli* | - | + | - | YES | YES | YES | no | 11186 |
| 432 | *E. coli* | - | - | - | YES | YES | no | no | 15407 |
| 437 | . | . | . | . | . | . | no | no | 14160 |
| 440 | *E. coli* | - | - | - | YES | YES | no | no | 10862 |
| 440 | *E. coli* | - | + | - | YES | YES | no | no | 12211 |
| 440 | . | . | . | . | . | . | no | no | 15722 |
| 441 | . | . | . | . | . | . | no | no | 14050 |

**Table S5. Classification of participants with diagnosed UTI by uropathogen and baseline fecal *E. coli* carriage**

| Days between Fecal sample and UTI | Uropathogen [a] | Uropathogen's Resistance to: | | | UTI sample available for clonal typing | Clonally matched Uropathogen *E. coli* found in fecal sample | ST131-*H*30 in fecal sample | ST1193 in fecal sample | Study ID |
|---|---|---|---|---|---|---|---|---|---|
| | | CIP | TMP/SXT | 3GC | | | | | |
| 442 | *E. coli* | - | - | - | no | . | no | no | 14918 |
| 447 | *K. pneumoniae* | - | - | - | no | . | no | no | 12041 |
| 451 | *K. pneumoniae* | - | - | - | no | . | no | no | 12805 |
| 452 | *E. coli* | - | - | - | YES | YES | no | no | 14871 |
| 456 | *E. coli* | - | - | - | YES | YES | no | no | 13064 |
| 456 | *P. mirabilis* | - | - | - | no | . | no | no | 13446 |
| 458 | *E. coli* | - | - | - | YES | YES | no | no | 15737 |
| 463 | *E. coli* | - | - | - | YES | no | no | no | 15808 |
| 463 | . | . | . | . | . | . | no | no | 15857 |
| 466 | . | . | . | . | . | . | no | no | 14844 |
| 467 | *Klebsiella sp.* | - | - | - | no | . | no | no | 15189 |
| 468 | . | . | . | . | . | . | no | no | 10919 |
| 468 | *E. coli* | - | - | - | YES | YES | no | no | 11172 |
| 471 | *E. coli* | - | - | - | YES | YES | no | no | 11620 |
| 472 | *E. coli* | + | - | - | YES | no | no | no | 13561 |
| 480 | *E. coli* | - | - | - | YES | no | no | no | 14030 |
| 481 | *E. coli* | - | - | - | YES | no | no | no | 12198 |
| 481 | *E. coli* | - | - | - | YES | no | no | no | 15855 |
| 483 | . | . | . | . | . | . | no | no | 16665 |
| 484 | *E. coli* | - | - | - | YES | no | no | no | 12748 |
| 488 | *E. coli* | - | - | - | YES | YES | no | no | 12096 |
| 493 | *Salmonella sp.* | - | - | - | no | . | no | no | 10944 |
| 493 | . | . | . | . | . | . | YES | no | 15280 |
| 494 | *E. coli* | - | - | - | YES | YES | no | no | 12244 |
| 500 | *E. coli* | - | - | - | YES | no | YES | no | 12681 |
| 503 | . | . | . | . | . | . | no | no | 10134 |
| 504 | *E. coli* | - | - | - | no | . | no | no | 15505 |
| 507 | *E. coli* | - | - | - | YES | no | no | no | 14421 |
| 508 | *E. coli* | - | - | - | YES | no | no | no | 12963 |
| 510 | *E. coli* | - | + | - | YES | YES | no | no | 11595 |
| 511 | *E. coli* | - | - | - | YES | no | no | no | 12255 |
| 514 | *E. coli* | - | + | - | YES | YES | no | no | 15034 |
| 514 | *E. coli* | + | - | - | YES | YES | no | no | 16663 |
| 516 | *E. coli* | + | - | - | YES | no | no | no | 10152 |
| 518 | . | . | . | . | . | . | no | no | 15033 |
| 521 | *E. coli* | + | + | - | YES | YES | no | YES | 16419 |
| 527 | *E. coli* | - | - | - | YES | YES | no | no | 14171 |
| 530 | *E. coli* | - | - | - | YES | no | no | no | 16637 |
| 531 | . | . | . | . | . | . | no | no | 11163 |
| 542 | *E. coli* | - | + | - | YES | YES | no | YES | 12118 |
| 542 | *E. coli* | - | + | - | YES | YES | no | no | 12815 |
| 558 | . | . | . | . | . | . | no | no | 14550 |
| 674 | . | . | . | . | . | . | no | no | 14497 |

[a] In 50/184 UTI cases uropathogen was not identified; missing data indicated by '.'.
